# Comparative analysis of the evolution of Life Expectancy in the United Republic of Tanzania, Uganda, and Kenya in 61 years (1960-2021): A secondary data analysis of the World Population Prospects (WPPs) on the three East African countries

**DOI:** 10.1101/2024.04.12.24305722

**Authors:** Gaye Bamba, Joelle Abi abboud, Emmanuel Olal, David Lagoro Kitara

## Abstract

**Background:** Life expectancy at birth (LE_0_) in Kenya, Uganda, and the United Republic of Tanzania in 1960 was 57, 54, and 42 years, respectively. However, in 2019, LE_0_ had gained in Kenya, Uganda, and Tanzania to 62.94, 62.99, and 66.99 years, respectively. This study aimed to evaluate and compare the progression of LE_0_ in Kenya, Uganda, and Tanzania over 61 years (1960-2021).

**Methods:** Life tables from World Population Prospects (WPPs) were used to calculate LE_0_ for Kenya, Uganda, and Tanzania by sex from 1960 to 2021. LE_0_ was contextualized alongside trends in 1960 and 2019. Using decomposition techniques, we examined how each sex contributed to losses or gains in LE_0_ between 1960 and 2021 and the likely contributory factors to LE_0_ losses. RStudio software was used to calculate differences in LE_0_ from one year to another. Linear regression analyses were used to trace the progression of LE_0_ in Kenya, Uganda, and Tanzania in six decades.

**Results:** LE_0_ improved from 1960 to 2021 in males and females in Kenya, Uganda, and Tanzania. The most substantial improvement occurred between 1960-1980 in Kenya and Tanzania while in Uganda between 1960-1970. LE_0_ losses were observed between 1980-2000, and 2020-2021 in Kenya and Tanzania while Uganda experienced losses between 1970-1981; 1989-1993, and 2020-2021. LE_0_ losses in Kenya, Uganda, and Tanzania were likely a result of deaths related to high infant, maternal, and child mortality rates due to infectious and non-communicable diseases (1990-2021). In Uganda, the political and economic turmoil during Amin’s regime (1971-1979) registered the most substantial LE_0_ losses over the period. In addition, LE_0_ gaps between males and females fluctuated over the years with the highest at 9.5 years in Uganda in 1982 and the lowest at 2.25 years in Kenya in 2001. The fluctuating LE_0_ gaps between males and females has been observed in the three East African countries.

**Conclusion:** LE_0_ in Kenya, Uganda, and Tanzania progressively increased from 1960-2021. Males and females showed fluctuating LE_0_ gaps in the last 61 years but females lived longer than males. There were LE_0_ losses between 1970-2000 and between 2020-2021. High infant, maternal, and child mortality rates, and later, the high prevalence of HIV and AIDS, immunizable diseases, and the COVID-19 pandemic were the likely reasons for LE_0_ losses. The COVID-19 pandemic contributed to LE_0_ losses more in Kenya than Tanzania and Uganda likely due to deaths related to the virus itself or the control measures.

LE_0_ losses in Uganda in the 70s were likely due to political and economic turmoil during the brutal Amin’s regime. Even though many studies show LE_0_ gains in Kenya, Uganda, and Tanzania over the 61 years, political and economic stability, economic growth, health systems strengthening, control of infectious diseases, and epidemics were critical in the LE_0_ gains. Thus, a more comprehensive study is warranted to assess the actual impact of public health interventions in LE_0_ gains in the three East African countries.

## Introduction

Globally, Life expectancy (LE_0_) has increased by almost 20 years over the last five decades.^1^ These increases have contributed hugely to gains in well-being worldwide^2^ and in the United States of America.^3^ Life expectancy (LE_0_) has long been hypothesized as an important determinant of individuals’ decisions to invest in physical and human capital (Becker, 1964; Ben-Porath, 1967) and this premise of economic theory has played a central role in the study of economic development.^4^

Given the potential importance of life expectancy in decision-making, there is growing recognition of the value of measuring individuals’ subjective expectations about future health and studying factors that influence these expectations.^5^ In the United States of America (USA) and other developed countries, elicitation of individuals’ expectations about survival to advanced ages and major life events has become fairly common.^6^ For example, the Health and Retirement Survey has included such measures^5^, and several studies have shown that eliciting individuals’ expectations is not only feasible but also useful for understanding important aspects of individuals’ decisions.^6^

In developing countries, however, very few population-level studies have elicited individuals’ expectations about longevity.^7^ Particularly in sub-Saharan Africa (SSA), which saw dramatic declines in life expectancy after the onset of HIV and AIDs epidemic followed by large reductions in adult mortality as HIV treatment with antiretroviral therapy (ART) was scaled up in the past decade.^8^ Also, there have been strikingly few studies examining associations between HIV and AIDS and individuals’ subjective expectations about life expectancy.^8^

The importance of health as an integral input in economic development is succinctly captured by the erstwhile Millennium Development Goals (MDGs) and now the Sustainable Development Goals (SDGs).^9^ In particular, life expectancy is one of the key indicators for assessing the level of a country’s progress as well as the efficacy of its health system.^9^

The sub-Saharan Africa has made progressive gains in life expectancy over the past two decades.^9^ For instance, between 1980 and 2000, life expectancy in sub-Saharan Africa increased from 46 to 56 years.^9^ However, its life expectancy lags behind the other regions of the world. For example, the Sierra Leone’s life expectancy of 47 years in 2010 was one of the lowest in the world.^9^

The UNDP (2006) states that life expectancy in sub-Saharan Africa was lower 30 years ago largely because of the devastating impact of HIV and AIDS.^10^ Thus, the global improvement in life expectancy notwithstanding, there exists a wide gap between life expectancy in the developed countries and that of the developing countries especially those in sub-Saharan Africa.^11^ For instance, countries such as Japan, Switzerland, and San Marino have a life expectancy of 83 years while over ten countries in SSA including Sierra Leone, Central African Republic, Guinea-Bissau, Somalia, Lesotho, and Mozambique have life expectancies between 47 to 53 years.^11^

Life expectancy (LE_0_) has increased in the majority of African countries for the period 1980-2021, with an average increase slightly higher than that of the world average for that period.^12^ Due to the low starting values of LE_0_ in 1960, and the slow increase in the period 1960-1980, many sub-Saharan countries presented in the year 2021, low values in comparison with the world average: 59 years in Sub-Saharan Africa and 71 as the World average.^12^ However, northern Africa has experienced a very positive evolution reaching the world average in the year 2014.^12^

In the East African countries of Uganda, Kenya, and the United Republic of Tanzania, LE_0_ gains have followed similar patterns with the rest of the sub-Saharan African countries however, there may be some in-country variations based on the history of each of the three East African countries over the years.

Amidst the sub-Saharan Africa’s best decade of economic growth since the 1970s, the East African Community (EAC) has been among the fastest-growing regions.^13^ Growth rates have picked up strongly since 2000, outpacing the rest of sub-Saharan Africa.^13^ During 2005–2011, per capita income growth reached 3.6% a year in the EAC, compared with 3.0% for sub-Saharan Africa as a whole, quadrupling the rate achieved in the previous fifteen years.^13^ Part of the recent high economic growth is a catch up after years of very poor growth in the last part of the 20th century, when the region suffered periods of severe civil strife and bouts of economic instability.^13^ Since then, Governments in EAC countries have been committed to strong economic policies.^13^

This study aimed to evaluate and compare the progression of life expectancy at birth in Kenya, Uganda, and the United Republic of Tanzania from 1960 to 2021 (61 years).

## Methodology

### Study design

We conducted a secondary data analysis on Kenya, Uganda, and the United Republic of Tanzania’s life expectancy from 1960 to 2021.

### Data

Life table data were categorized by age, country, and by sex (female, male, and both). They were extracted from the World Population Prospects (WPPs) estimate for 2022.^14,15^ From these life tables, we extracted data for Kenya, Uganda, and the United Republic of Tanzania. Too, we extracted data from 1960 to the latest available year at the time of writing (2022).^14,15^ All life tables were downloaded on March 28^th^, 2023. These life tables on WPPs offer detailed information on both sexes, including single-age mortality rates up to the age of 100.^14,15^ It presents a comprehensive set of values, showcasing the mortality experience of a hypothetical cohort of infants born simultaneously and subjected to the specific mortality rates of a given year throughout their lifetime.^14,15^

The table encompasses essential metrics, such as probabilities of dying (qx) and surviving (px), counts of individuals surviving (lx) and dying (dx), total person-years lived (Lx), survivorship ratios (Sx), cumulative stationary population (Tx), average remaining life expectancy (ex), and average number of years lived (ax).^14,15^ In addition, we added a column for sex, including female, male, and both, to combine the two tables into a unified table containing data from 1960 to 2021 for the three countries.^14,15^

### Life expectancy values

In this study, we utilized life expectancy (LE_0_) measures from life tables to observe the progression of life expectancy at birth over 61 years (1960 to 2021) in Kenya, Uganda, and Tanzania. Additionally, we focused on life expectancy at age 0 (LE_0_) as it is one of the most commonly used metrics for determining population health and longevity.^15^ We calculated differences in LE_0_ to compare life expectancy progression in the three countries, years, and sexes.^15^

### Statistical analysis

For descriptive analysis, the life table was divided into two categories: one for males, and another for females. These categories were then used to characterize life expectancy at birth (LE_0_) using means and standard deviations (Sd). A linear regression model, considering life expectancy as a continuous variable, was used to estimate the relationship between age, years, sex, and life expectancy in Kenya, Uganda, and Tanzania. In addition, a sex-stratified analysis was conducted to examine the impact of gender on life expectancy.

Furthermore, a separate set of correlations and analysis of variance were performed to evaluate the relationship between years and age with the life expectancy of the three East African countries. All statistical analyses were conducted using RStudio version 4.2.2.^16,17^

### Yearly changes in life expectancy at birth (LE_0_) in Kenya, Uganda, and Tanzania

In this analysis, we employed the annual changes in life expectancy (LE), which was calculated as the difference between LE in a given year (LE□) and the previous year (LE□_−1_), where i represented the year.

### Linear Regression Analyses

The formulae below were used to calculate life expectancy (LE_0_) in Kenya, Uganda, and Tanzania over 61 years (1960-2021).

Differences in life expectancy (LE_0_) between females and males=β_0_+β_1·_Year+β_2·_Countries+ε (1).

The formula for the linear regression is ex=β_0_+β_1·_Year+β_2·_age+β_3·_sex+ε (2).

In the calculation, LE_0_ represents the predicted value of the dependent variable (Life expectancy at birth). β_0_ is the intercept, representing the value of LE_0_ when all predictor variables are zero. β_1_ is the coefficient for the “Year” predictor variable, indicating how much the predicted value changes for a unit change in the “Year” variable while keeping other predictors constant. β_2_ is the coefficient for the “Age” predictor variable, indicating how much the predicted value changes for a unit change in the “Age” variable while keeping other predictors constant. β_3_ is the coefficient for the “Sex” predictor variable, indicating how much the predicted value changes when the “Sex” variable changes (assuming it’s a categorical variable, encoded as 0 or 1 for male/female). β_4_ is the coefficient for the “Three East African countries” predictor variable, indicating how much the predicted value changes for a unit change in the “country” variable while keeping other predictors constant.

## Results

There have been progressive Life expectancy gains in all three East African countries (Kenya, Uganda, and the United Republic of Tanzania) over the last 61 years (1960-2021). Uganda and Tanzania had lower LE_0_ and probability of survival in 1960 compared to Kenya. However, by 2021, Tanzania had the highest LE_0_ followed by Uganda and then Kenya even though the probability of survival was highest among Kenyans followed by Ugandans, and least among Tanzanians.

### Yearly changes in life expectancy at birth in the three East African countries (Kenya, Uganda, and the United Republic of Tanzania) from 1960 to 2021

In Tanzania, life expectancy at birth has exhibited gradual gains from 1960 to 1987, followed by a period of decline from 1988 to 1998. From 1999 to 2019, there has been a steady increase in LE_0_ from one year to the next (Figure 3). However, with the onset of the COVID-19 pandemic in 2020, LE_0_ experienced a notable decline compared to 2019 (−0.58 years). This decline continued into 2021 but to a lesser extent than in 2020, with a decrease of 0.21 years (Figure 3).

In Kenya, LE_0_ increased compared to the previous year from 1960 to 1983, with exceptions in 1974 and 1982 when LE_0_ decreased compared to the preceding years (Figure 3). From 1984 to 2000, there was a continuous decline in LE_0_. However, starting in 2001, there was a progressive increase in LE_0_ until 2019. With the COVID-19 pandemic in 2020, LE_0_ decreased compared to 2019 (−0.26 years). The decrease was even more pronounced in 2021 compared to 2020, with a loss of more than -1 year of LE_0_ (−1.26 years). Kenya has experienced more years with losses in LE_0_ compared to Tanzania. This may explain why LE_0_ in Tanzania has surpassed Kenya in the year 2021 (Figure 3).

Uganda’s life expectancy pattern has substantially varied with the other two East African countries (Kenya and Tanzania). Its LE_0_ increased from 1960 to 1970. From 1971 to 1981 LE_0_ in Uganda experienced progressive losses but gains were experienced again from 1982 to 1986. Thereafter life expectancy losses were observed from 1988 to 1993. From 1994 to 2019 there were consistent LE_0_ gains in Uganda (Figure 3). The emergence of COVID-19 from 2020 to 2021 cut short the progressive LE_0_ gains and posted losses in the two years (Figure 3).

### Linear Regression Analyses

Linear regression analyses were used to trace the progression of life expectancy in Kenya, Uganda, and the United Republic of Tanzania in the last six decades. The findings are that Kenya’s LE_0_ gains from 1960 to 1964 (LE, p=0.1); from 1965 to 1976 (p<0.001); from 1997 to 2004 (p=0.1); and from 2005 to 2021 (p<0.001) (Table 1 and Table 2).

In Uganda, LE_0_ gains were: from 1962 to 1970 (p<0.001); from 1971 to 1979 (p=0.1); from 1980 to 2005 (p<0.001); from 2006 to 2011 (p=0.1) and from 2012 to 2021 (p<0.001) (Table 3 and Table 4).

In the United Republic of Tanzania, LE_0_ gains patterns were as follows; from 1961 to 1966 (p=0.1) and from 1967 to 2021 (p<0.001) (Table 5 and Table 6).

## Discussion

This study observed progressive life expectancy gains in all three East African countries (Kenya, Uganda, and the United Republic of Tanzania) over the last 61 years (1960-2021) (Figure 1, Figure 2, Figure 3, Figure 4, Figure 5, Figure 6). Even among different ages and sexes, there is evidence of LE_0_ gains in all three East African countries although with different patterns (Figure 7, and Figure 8). Uganda and Tanzania had lower LE_0_ and probability of survival in 1960 compared to Kenya (Figure 9). However, by 2021, Tanzania had the highest LE_0_ followed by Uganda and then Kenya even though the probability of survival was highest among Kenyans followed by Ugandans, and least among Tanzanians (Figure 9). These findings were consistent with LE_0_ gains observed in most sub-Saharan Africa (SSA) countries and globally over the years.^1-11^

**Figure 1:**
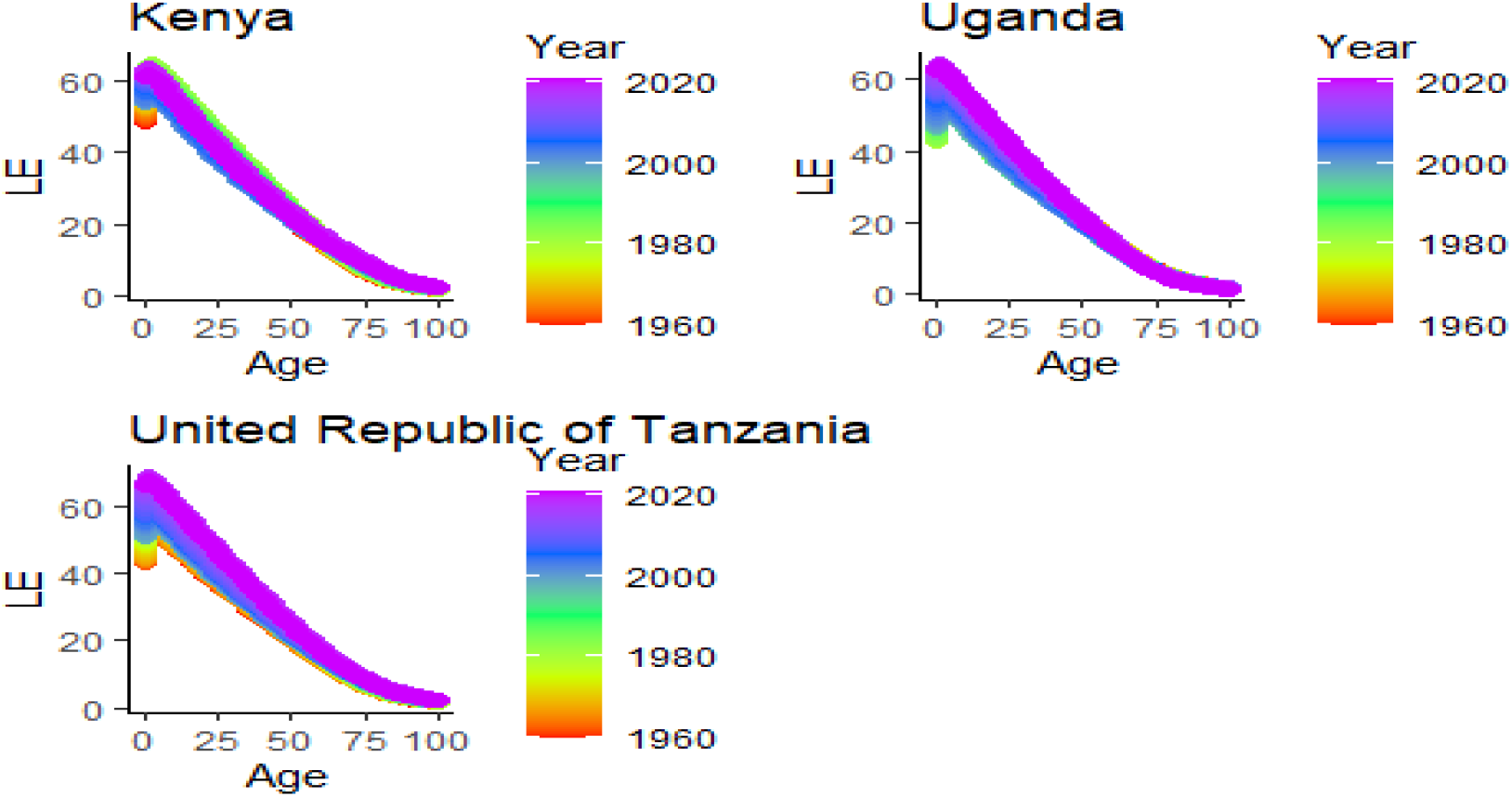
Life expectancy variations by years in Kenya, Uganda, and the United Republic of Tanzania over 61 years (1960-2021). These graphs show visual variations in the evolution of life expectancy over the years for different age groups In Kenya, Uganda, and the United Republic of Tanzania. From these graphs, it is evident that LE_0_ in Tanzania and Uganda have gradually increased over the years across all age groups. Also, in Kenya, life expectancy at age 0 and 60 and above have shown gradual increases over the years. However, for ages 1 to 59, it is noticeable that life expectancy was higher from 1969 to 1992, whereas in recent years, there has been a decline in life expectancy in this age group. This decline is observed in males and females.

**Figure 2:**
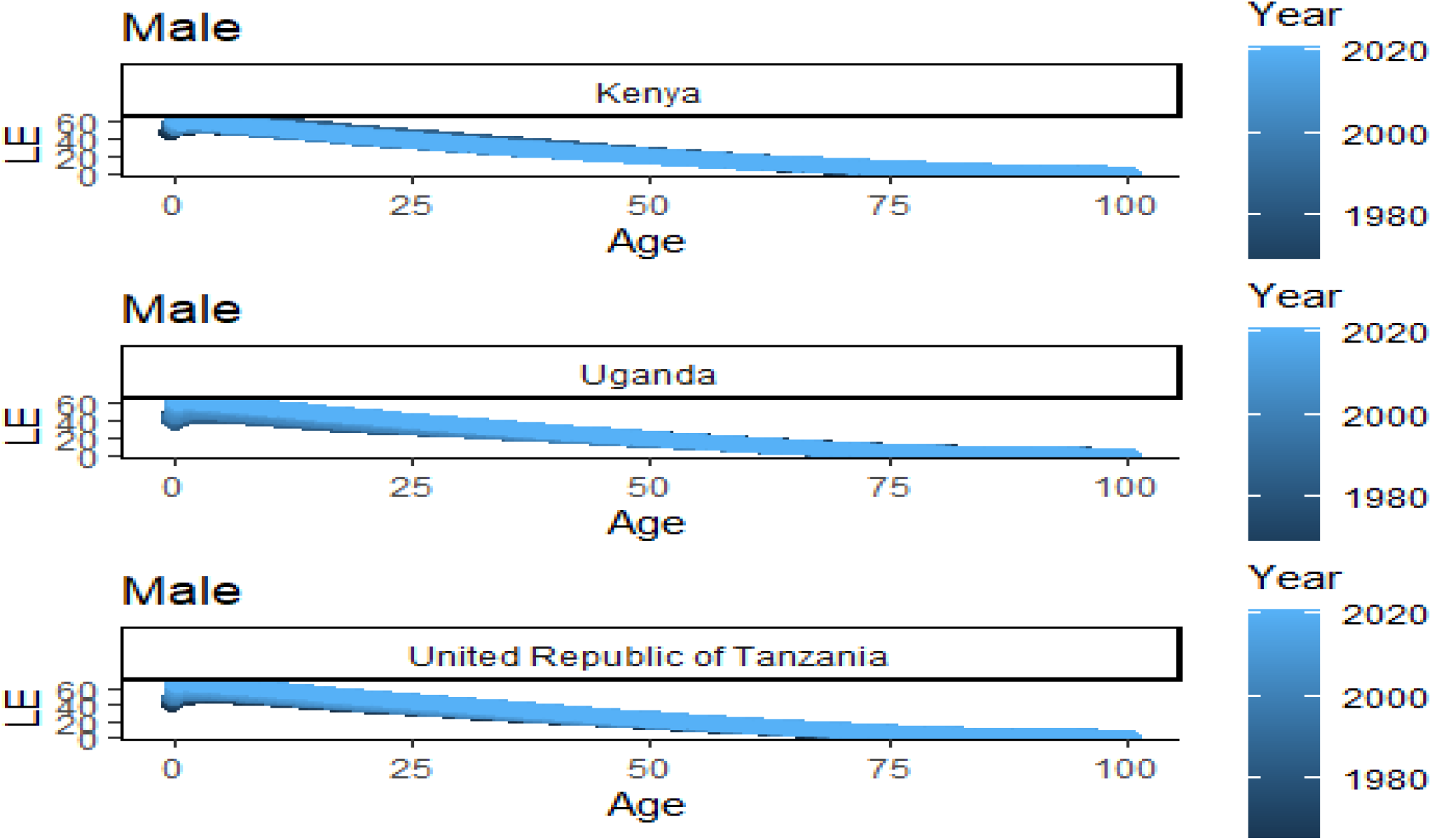
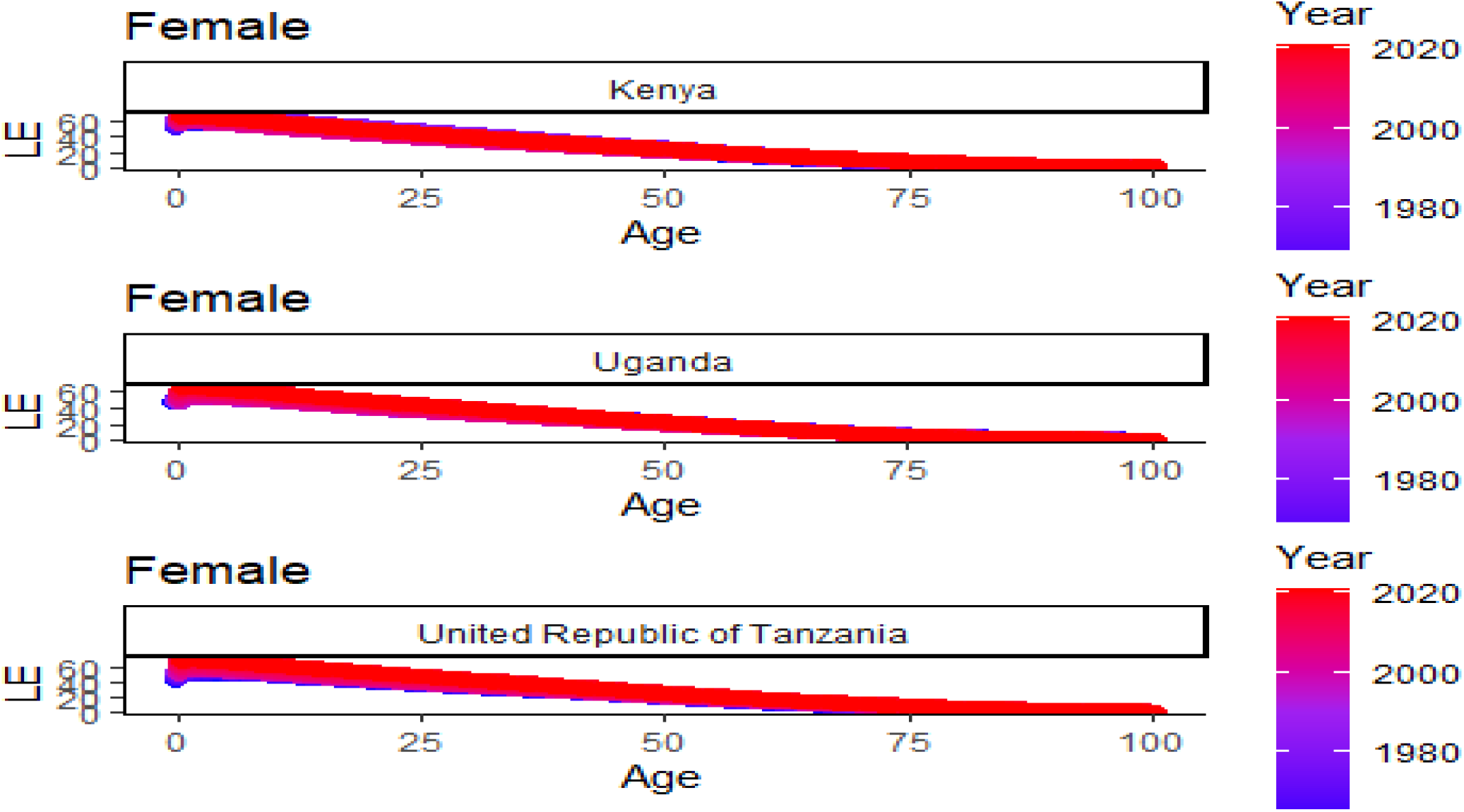
Life expectancy variations by years among males and females in Kenya, Uganda, and the United Republic of Tanzania over 61 years (1960-2021). The figure displays the gradual drop in life expectancy among males and females in Kenya, Uganda, and the United Republic of Tanzania at increasing ages of 25, 50, 75 and 100 years. The pattern in Kenya differs to that of the United Republic of Tanzania and Uganda whereas the Kenyan cases where declining faster at the 50-year mark in both sexes.

**Figure 3:**
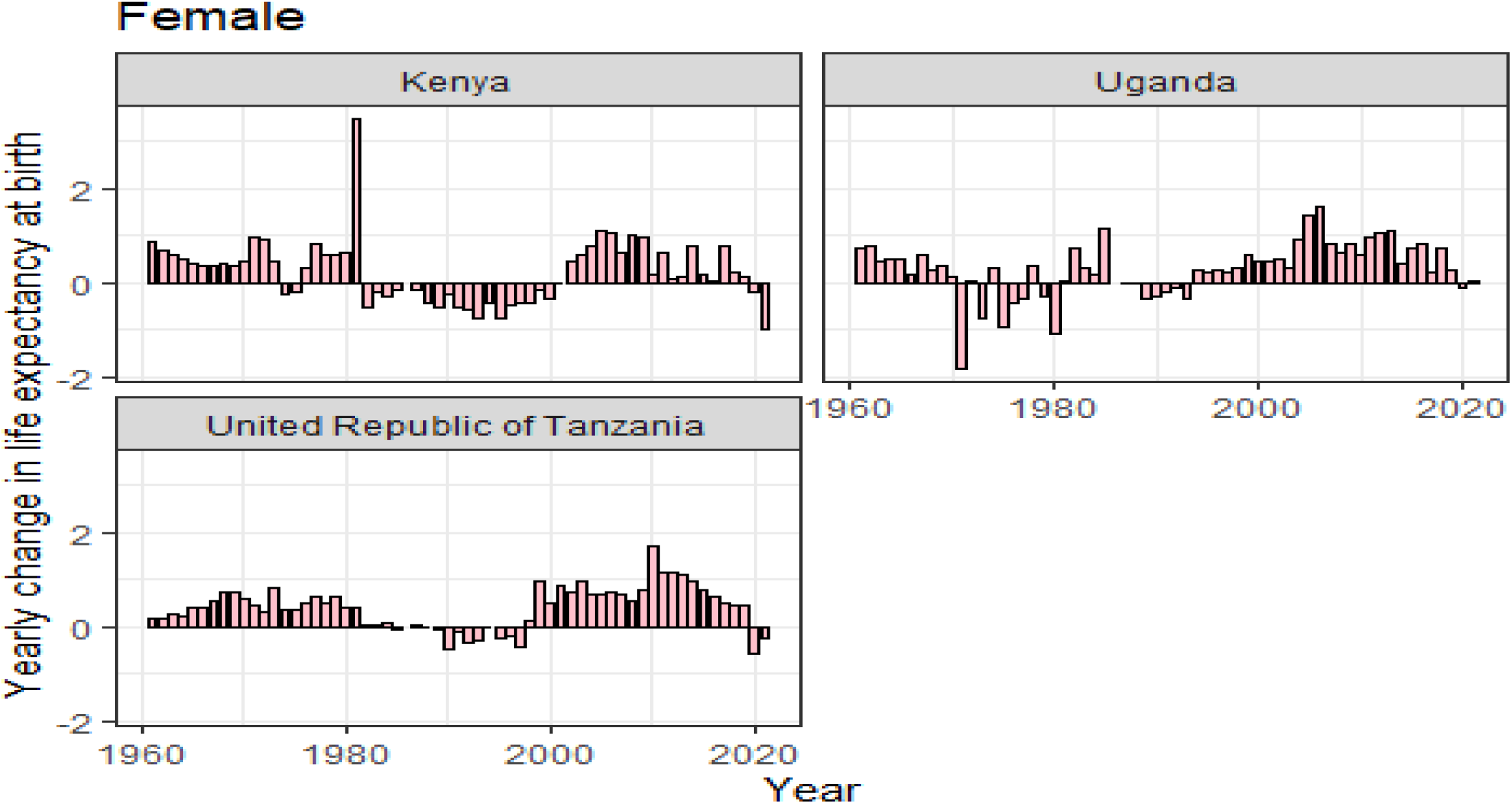
Life expectancy changes in Kenya, Uganda, and United Republic over 61 years (1960-2021). The figure shows that in Kenya, life expectancy gains were observed between 1960 to 1981 however, losses were observed between 1982 to 2000 and also 2020 to 2021. In Uganda, life expectancy gains were observed between 1960 to 1970 and losses were observed between 1971 to 1980 and also between 2020 to 2021. In the United Republic of Tanzania, life expectancy gains were observed between 1960 to 1985 but losses were observed between 1989 to 1998 and 2020 to 2021.

**Figure 4:**
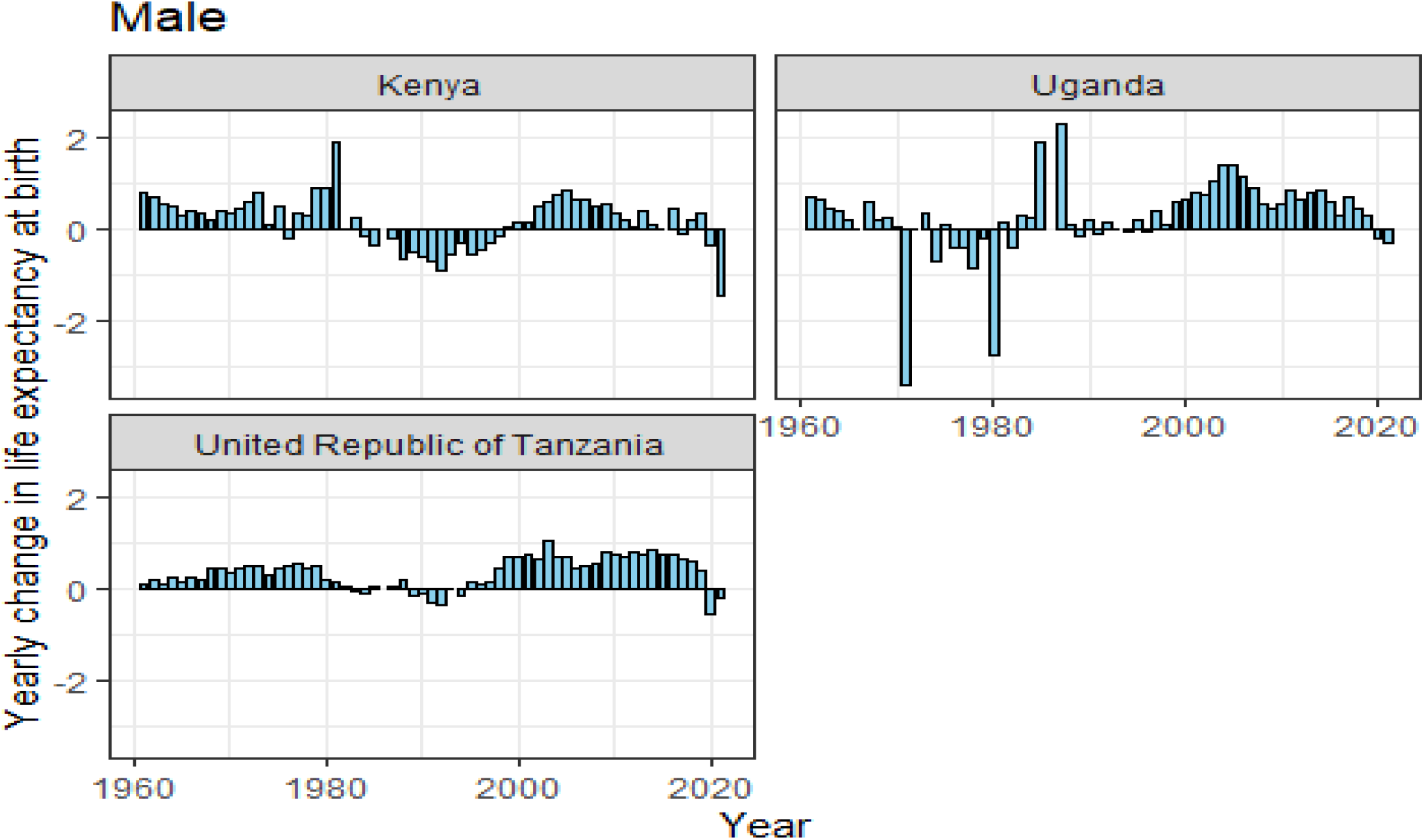
Life expectancy changes among females in Kenya, Uganda, and The United Republic over 61 years (1960-2021). The figure shows that in Kenya, life expectancy gains among females were observed between 1960 to 1973, then 1975 to1982, however, losses were observed between 1974-1975; 1982-2000; and 2020-2021. In Uganda, life expectancy gains in females were between 1960-1970 but losses were observed between 1971-1981 and between 1988-1993, and in 2020. In the United Republic of Tanzania, life expectancy gains among females were observed between 1960-1985 but losses were observed between 1986-1997 and 2020-2021.

**Figure 5:**
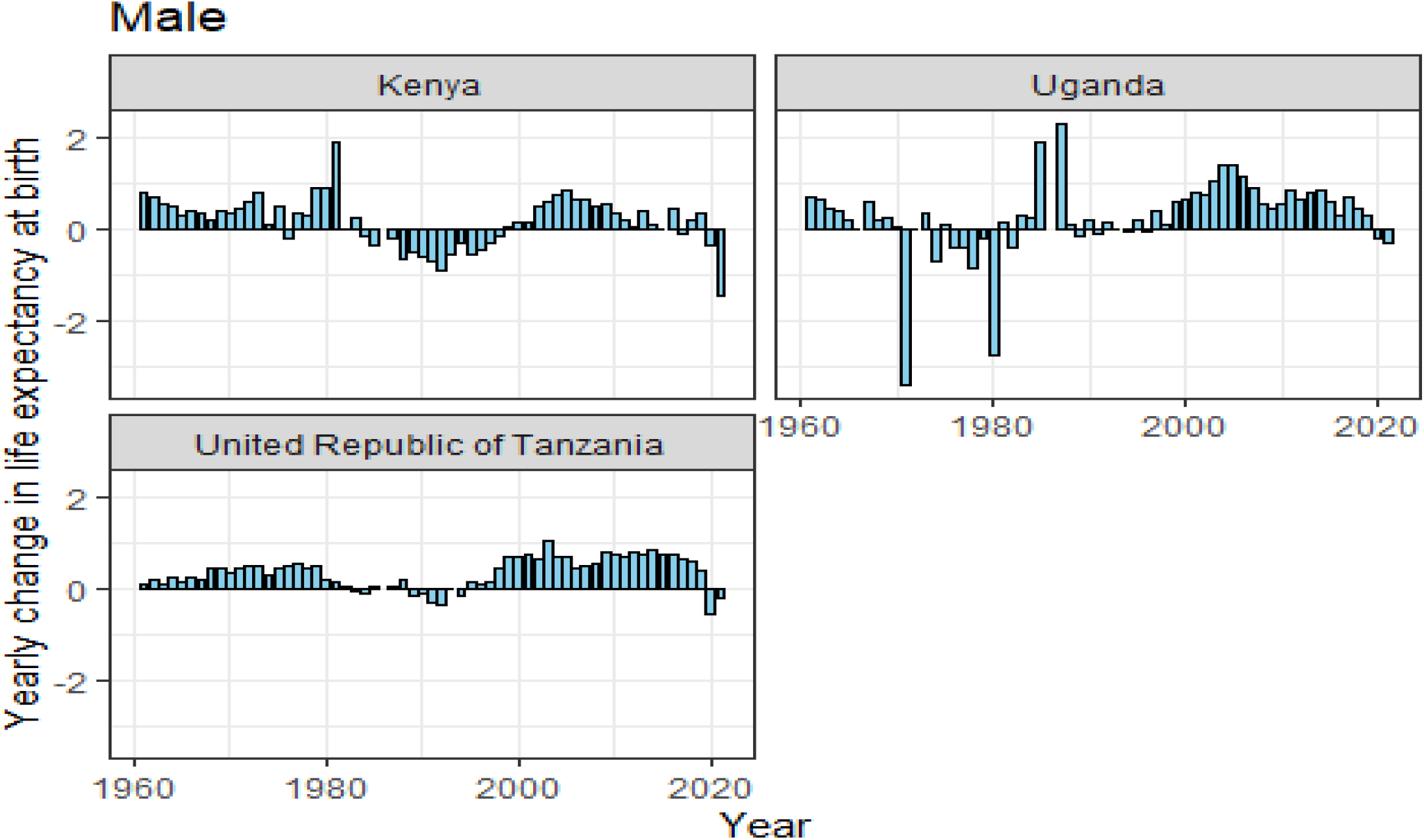
Life expectancy changes among males in Kenya, Uganda, and The United Republic over 61 years (1960-2021). The figure shows that in Kenya, life expectancy gains among males were observed between 1960-1981 however, losses were observed in 1976, and between 1983-1998; and 2020-2021. In Uganda, life expectancy gains among males were observed between 1960-1970 however, losses were observed between 1971-1982; 1989-1991 and 2020-2021. In the United Republic of Tanzania, life expectancy gains among males were observed between 1960-1983 but losses were observed between 1983-1993 and 2020-2021.

**Figure 6:**
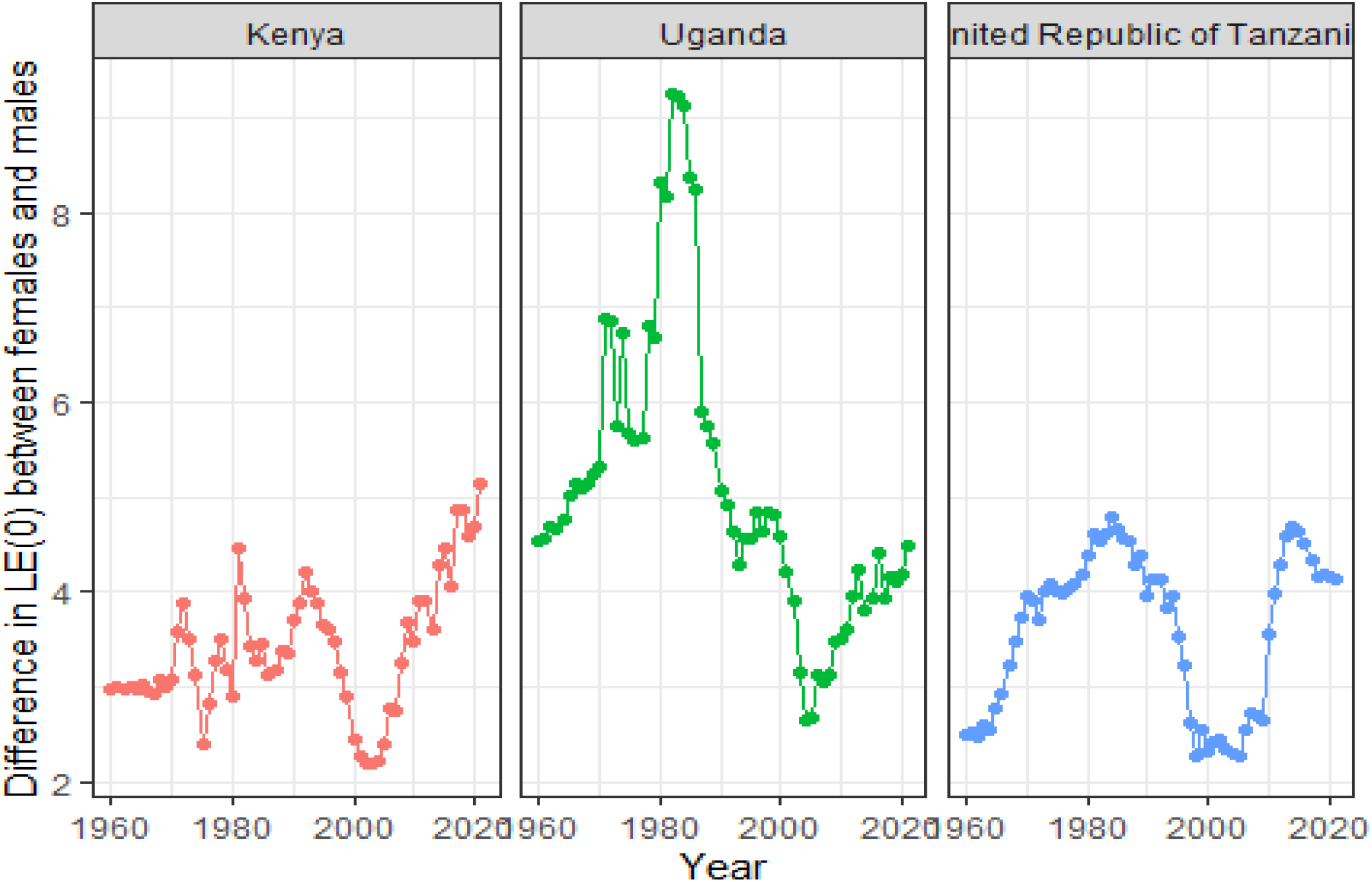
Gaps in life expectancy between males and females in Kenya, Uganda, and The United Republic of Tanzania over 61 years (1960-2021). In Kenya, the gap in life expectancy (LE_0_) at birth between males and females was 3 years in 1960, with females having a higher LE_0_ than males. This difference fluctuated over time but remained close to 3 years. In 1981, the gap increased to 4.5 years before it declined to 3.25 in 1986. It again rose to 4.25 in1992. From 1992, the gap declined, reaching its lowest point in 2002 and 2003, when the gap was only 2.20 years. Subsequently, the gap began to increase again, reaching 5.15 years by 2021. In Uganda, the gap in LE_0_ between males and females was 4.5 years in 1960, with females having a higher LE_0_ than males. This gap increased over time and reached the highest at 9.5 years in 1981. From 1981, the gap progressively declined to the lowest point at 2.5 years in 2005 but from then it increased to 4.5 years in 2021. In United Republic of Tanzania, the gap in LE_0_ between males and females was 2.5 years in 1960, with females having a higher LE_0_ than males. This gap increased over time, reaching the highest at 4.78 years in 1984. From 1984 to 2005, the gap declined reaching its lowest point of 2.28 years in 2005. Subsequently, the gap started increasing again, reaching 4.68 years by 2014. However, from 2014 to 2021, the gap declined and reached 4.13 years in 2021.

**Figure 7:**
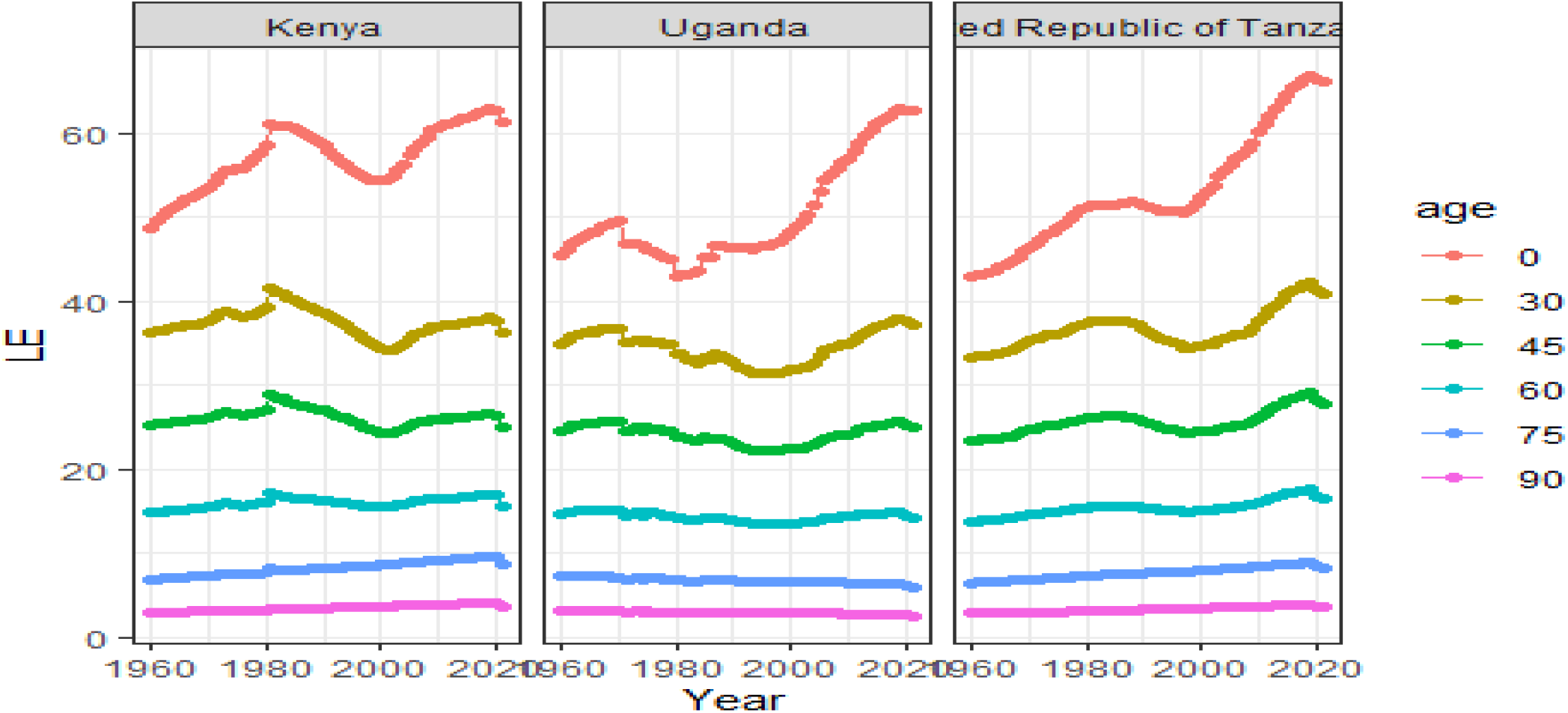
Life Expectancy (LE_0_) changes in Kenya, Uganda, and the United Republic of Tanzania from 1960 to 2021 in different ages. Figure 7 shows that the highest life expectancy at birth (LE_0_) for the three East African countries occurred in 2019, with Tanzania achieving the highest than Kenya and Uganda. Specifically, life expectancy at birth was 62.94 years for Kenya, 62.99 years for Uganda, and 66.99 years for Tanzania. Following 2019, the second-highest LE_0_ for both countries was observed in 2018, with the year 2020 ranking third. It’s noteworthy that in 1960, life expectancy at birth in Kenya was higher than in Uganda and Tanzania. However, the consistent increase in life expectancy in Uganda and Tanzania have been greater than Kenya over time.

**Figure 8:**
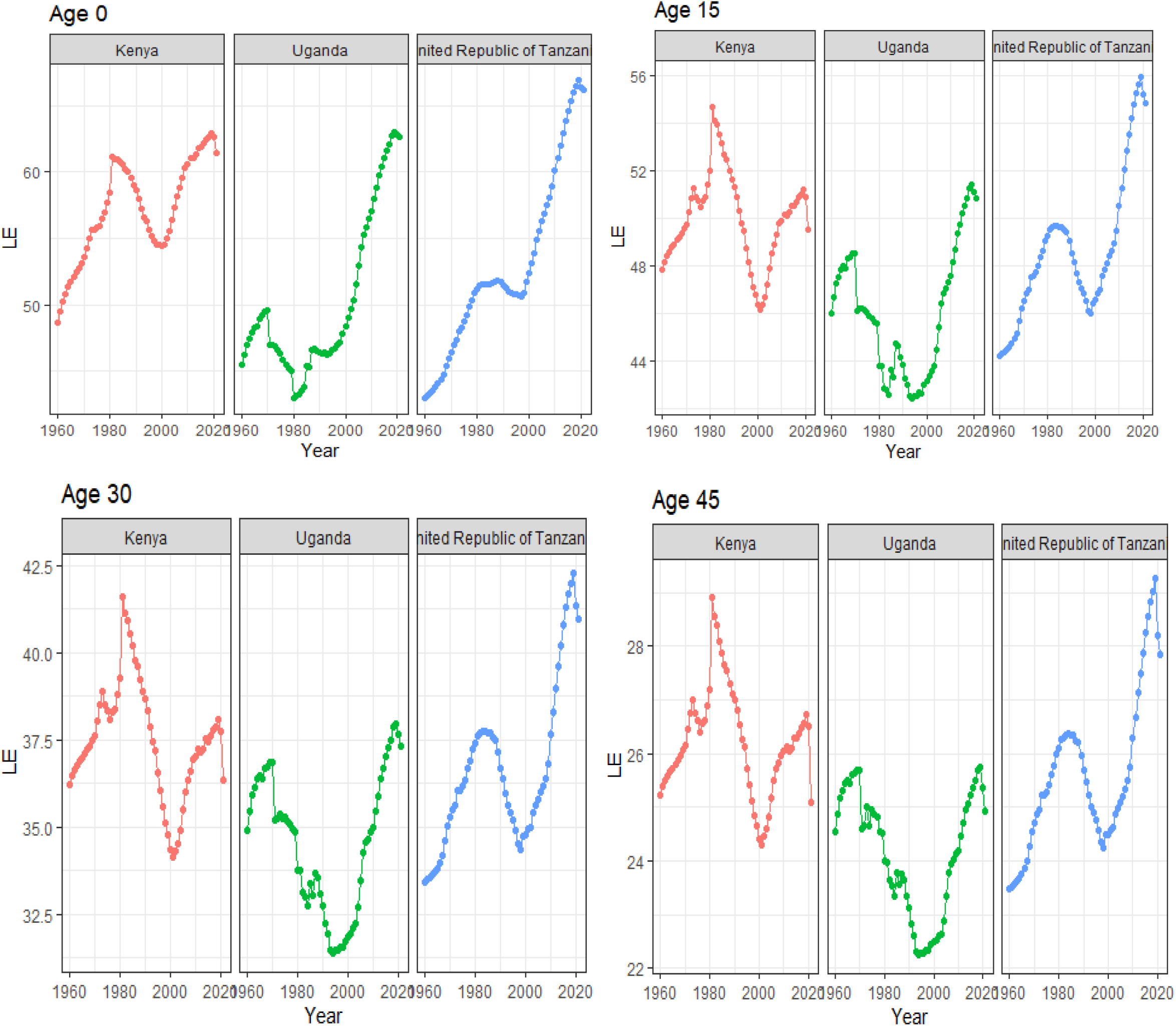
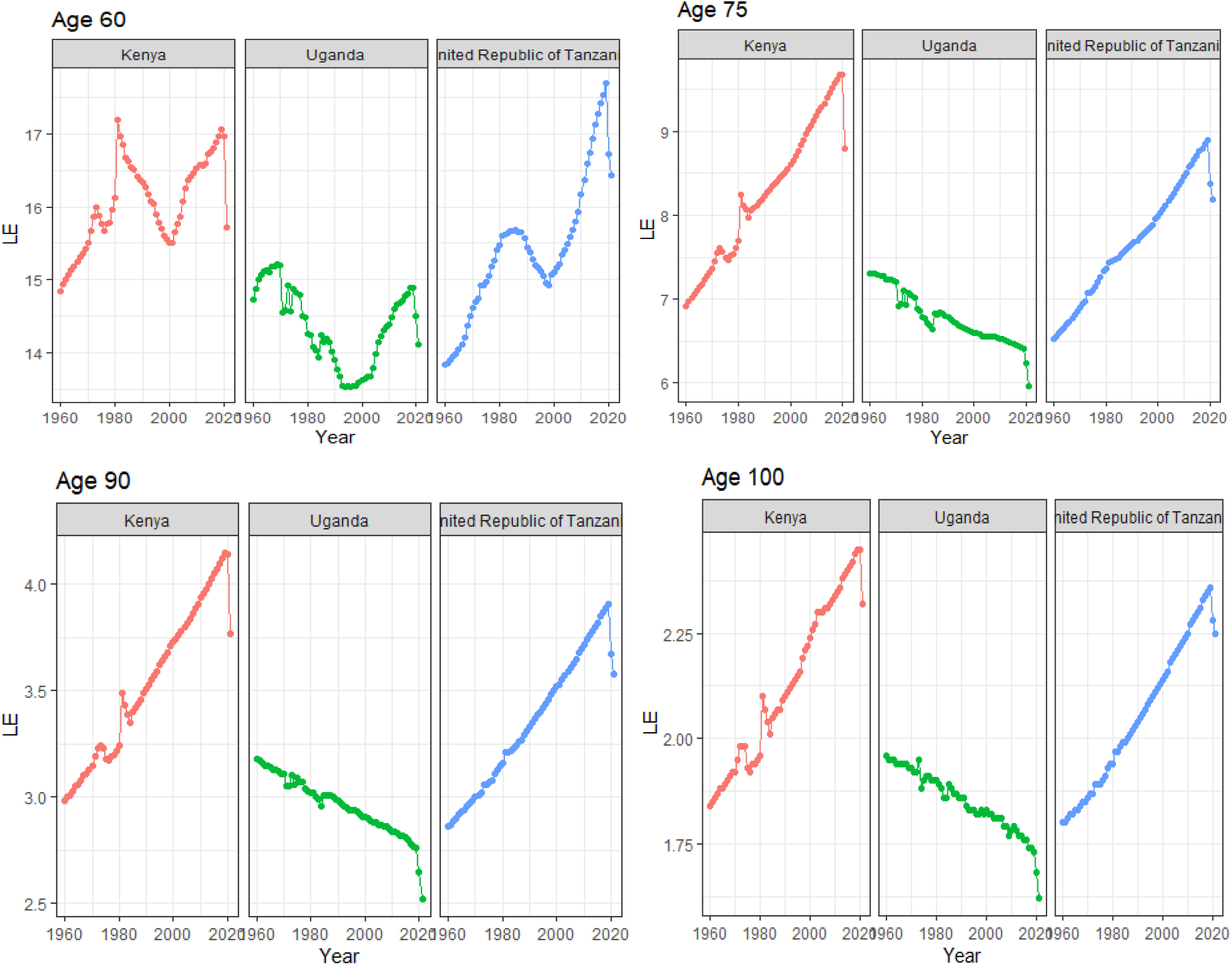
Life Expectancy (LE_0_) changes in Kenya, Uganda, and the United Republic of Tanzania from 1960 to 2021. The highest life expectancy at birth (LE_0_) for the three East African countries occurred in 2019, with Tanzania achieving the highest LE_0_ than Kenya and Uganda. Specifically, LE_0_ was 62.94 years for Kenya, 62.99 years in Uganda, and 66.99 years for Tanzania. Following 2019, the second-highest LE_0_ for both countries was observed in 2018, with the year 2020 ranking third. It’s noteworthy that in 1960, LE_0_ in Kenya was higher than Uganda and Tanzania. However, the consistent and larger increases in LE_0_ in Uganda and Tanzania over time led to higher LE_0_ than Kenya.

**Figure 9:**
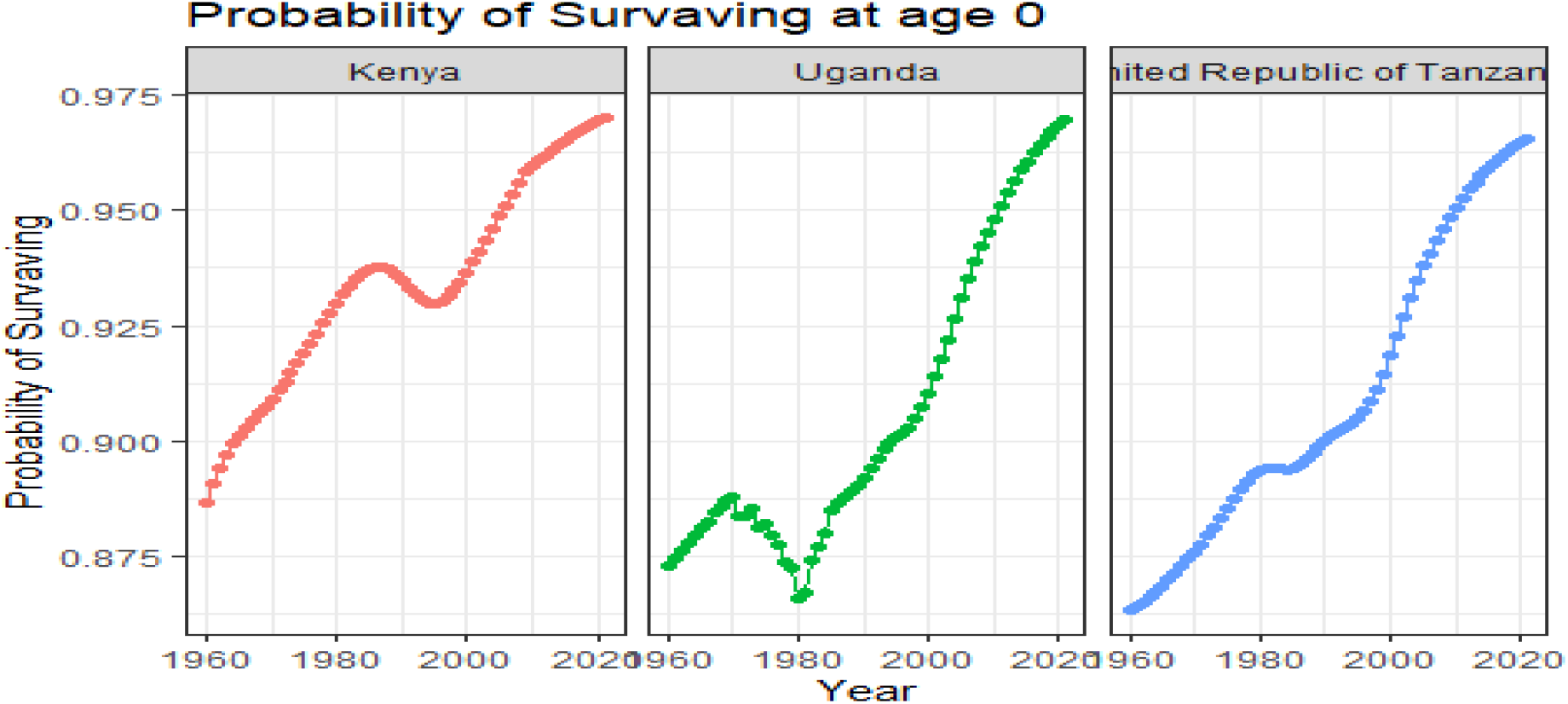
The probability of survival in the three East African Countries over 61 years. The probability of survival at birth increased in all three African countries (Kenya, Uganda, and the United Republic of Tanzania) from 1960 to 2021. Uganda and Tanzania had lower probability of survival in 1960 at 0.875 and 0.8625, respectively. In Kenya, the probability of survival was highest of the three East African countries in 1960 at 0.8875, and in 2021, at 0.965. In Uganda, it was 0.875 in 1960 and 0.8625 in 1980 which was the lowest probability of survival in Uganda over the last 61 years. In 2021, all three East African countries had a high probability of survival with the highest being in Kenya, followed by Uganda, and the least in the United Republic of Tanzania.

The WHO statistics of 2021 show an overall increase in global life expectancy and healthy life expectancy at birth as a result of improvements in the prevention and control of communicable diseases, maternal, perinatal, and nutritional conditions, non-communicable diseases, injuries, and their underlying determinants.^18^ Persisting inequalities also continue to impact population health in most, if not all, aspects.^18^ Despite the overall improvement in service coverage, between and within countries disadvantaged populations still have lower access to care and are at greater risk of facing catastrophic costs.^18^

While premature deaths from non-communicable diseases, the world’s leading cause of death continue to fall, progress has slowed in recent years and key risk factors including tobacco use and alcohol consumption, hypertension, obesity, and physical inactivity will require urgent and targeted interventions.^18^ Deaths from communicable diseases have also declined but continue to claim millions of lives each year, particularly in lower-resource settings where many people cannot access quality health services.^18^

Consistent with our findings, women live longer than men over the study period although there has been a progressive narrowing of the LE_0_ gap over the years (Figure 2, Figure 4, Figure 5, Figure 6). There has been a steady decrease in mortality from suicide, homicide, unintentional poisoning, and road traffic injuries, but many more of these deaths can still be prevented and men are at higher risk of dying from these causes than women.^18^ It is suggested that identifying health inequalities and their determinants is essential for achieving health equity and improving program delivery.^18^

Thus, knowing who is being left behind relies on equity-oriented national health information systems to produce and use inequality data for a fairer, and healthier world.^18^ Yet, high-quality disaggregated data for monitoring health inequalities and for ensuring equitable health service access and uptake are lacking worldwide, especially in sub-Saharan Africa.^18^ In addition, even the available disaggregated data are often not made accessible to decision-makers as needed.^18^ Notably, only 51% of 133 studied countries include data disaggregation in published national health statistical reports, ranging from 63% in high-income countries (HICs) to only 46–50% for other income groups.^18^

Thus, investments and political commitments are vital for enhancing country health information systems that generate disaggregated data by multiple inequality dimensions through various data sources including civil registration and vital statistics, population-based surveys, routine health facility data, and administrative data.^18^

The commitment to leave no one behind is a cornerstone of the 2030 Agenda for Sustainable Development. But within and between countries, high and rising inequalities act as both visible and concealed impediments to progress in population health and human, social, and economic developments.^18^

Thus, reducing inequality is a discrete SDG (SDG 10) and is vital to achieving all SDGs including ending poverty (SDG 1), ending hunger (SDG 2), ensuring healthy lives (SDG 3), ensuring inclusive and equitable quality education (SDG 4) and achieving gender equality (SDG 5).^18^

### Closing the Gender Gap

It is evident from our findings in East Africa that women are the healthier sex and real champions in the final game of life and longevity.^19^ They have an innate biological advantage over men, with female superiority in life expectancy appearing to be one of the most striking features of human biology.^19^ However, biology is only a part of the story, since it cannot answer why the gender gap in life expectancy would fluctuate over time.^19^ Biology and society interact with each other to produce gender-specific variations in diseases and mortality patterns.^19^ Thus, the relative influence of biology and society on the female advantage in life expectancy is an issue of debate.^19^

The biological gap in life expectancy between women and men appears to be a natural destiny that no fair society can avoid.^19^ On the contrary, the social gap in life expectancy is systematic and unjust.^19^ It is now known that we can rarely change our genetic and biological make-ups and thus cannot in principle eliminate the gender gap in longevity.^19^ Even if biology accounts for a small part of the male’s disadvantaged life span, gender equality in health may remain elusive.^19^ Thus, we can certainly narrow the gap by promoting healthy lifestyles and designing a society where both men and women have a fair chance to maximize their health potential.^19^

Consistent with our study findings, life expectancy improvements in sub-Saharan Africa vary between men and women.^19^ People in sub-Saharan Africa are now living longer than ever before but women live longer than men.^19^ A child born in the region today is expected to live up to 64 years on average.^19^ This is a remarkable increase of eleven years since the year 2000 when life expectancy at birth was only 53 years in sub-Saharan Africa.^19^

Therefore, monitoring life expectancy provides crucial information that’s needed to deploy resources and effective interventions on the ground.^19^

Elsewhere in the world, women have outlived men for more than a century as observed in the U.S. Demographers’ report largely attributed this well-known statistical gap to differences in behaviors in areas such as smoking and drinking habits, risk of injury, and drug use.^20^ Previous research reports have attributed biological factors such as more robust immune systems, to women’s relative longevity.^20^

In 2010 women were projected to live 4.8 years longer than men and in 2021 this gap widened to 5.8 years, the largest disparity since 1996.^21,22^ During the 20th century, heart disease was the main cause of death that created the difference in life expectancy between women and men.^21,22^ But in 2021, COVID fatalities and a growing number of drug overdoses among men were to blame according to a new analysis of CDC data published in JAMMA internal Medicine.^21,22^

A greater number of COVID deaths among men, for example, were influenced by a higher burden of comorbidities, differences in health behaviors, and employment in higher-risk industries which was a big part of the deeply troubling drop in life expectancy and widening gender gap.^21,22^ However, this report underscores some underlying public health problems that disproportionately affect men, especially drug overdoses, suicide, and other violence.^21,22^ Men are on average taller, more muscular, and seemingly stronger than women, and hence they are typically considered to be the stronger sex.^21,22^ Medical science, however, has a different story to tell us: men are biologically weaker than women.^21,22^ In almost all countries across the world, women outlive men^23^ and male mortality is higher for almost all primary causes of death.^24^ The female superiority in longevity is true not only for humans but also for many other species of mammals.^25,26,27^

Although women live longer than men, paradoxically they tend to report poorer health and more frequent hospital visits.^24^ Diseases experienced by men and women differ in terms of their prevalence and severity.^24^ There is, however, no conclusive evidence in support of the hypothesis that women are sicker than men.^24^ It has been observed that women consistently suffer from increased morbidity in the domain of psychological health measures, but no increased risk is found with physical health conditions.^24^

Women usually experience structural disadvantages in various spheres of society, which limits their potential to maximize health and well-being. Yet they paradoxically seem to be the healthier sex. The mechanisms that underlie the gender gap in health and mortality are complex and are not fully understood although several biological and social mechanisms can be suggested as explanations.^24^

### Biological mechanisms

From a biological point of view, men are naturally programmed to die earlier than women at the very moment of conception.^28,29^ Available evidence indicates that the male fetus is biologically weaker and more vulnerable to maternal stress and pregnancy complications than the female fetus.^28,29^ This is evident in the proportion of preterm births and the neonatal and infant mortality rates, which are higher in boys compared to girls.^30^ The sex differences at birth provide the foundation for the biological explanation of male disadvantage in life expectancy.^30,31,32^

As observed in the three East African countries, there has been a huge decrease in mortality among children younger than five in sub-Saharan Africa.^33^ As a portion of total deaths, the number of deaths before the age of five has decreased from 45% in 1950 to 10% in 2017.^33^ This was likely linked to several interventions such as scale-up of vaccination programs, improved water and sanitation, and mass distribution of insecticide-treated bed nets.^33^ In addition, mothers’ increased levels of education, economic growth, and rising individual incomes have also contributed to the decrease in children’s deaths.^33^ However, caution is necessary as it is inevitable that death rates will keep falling and the rising epidemics of high blood pressure, high blood sugar, and obesity in some African countries could lead to shifts over time in the opposite direction.^33^

### Falling death rates

As observed in the LE_0_ gains in the East African countries, the overall death rates have been dropping in many sub-Saharan African countries since the beginning of the 21^st^ century.^34^ It is reported that there were 702 deaths per 100,000 people in 2017, down from 1,366 deaths per 100,000 in 2000.^34^ This shows a tremendous improvement in mortality rates across sub-Saharan Africa. For all ages, it is estimated that one-third of life expectancy improvements are because of rising income per capita, one-third can be attributed to improvements in educational attainment, and one-third are a result of changes that happened over time.^12,13^ These include technological improvements such as new vaccines, and more information on diseases and how to control them. Additionally, there may be other reasons for the reduction in mortality over time which include social factors like the availability of job opportunities and good working conditions, the existence of social support networks, and safe housing.^12,13,34^

### Strengths and Limitations of the study

This study has many strengths in that the data used for this study was derived from live births and deaths across the world from the United Nations World population prospects. The information presented is reliable and trustworthy. In addition, regular updates of information are obtainable from the same database. However, this information obtained may not be detailed in subnational populations as these are aggregated datasets. In addition, countries that do not regularly or properly document birth and death rates may be under or overrepresented in the datasets due to the use of population estimates.

### Generalizability of findings

This information on the three East African countries (Uganda, Kenya, and the United Republic of Tanzania) can be generalized in the context of a developing country in sub-Saharan Africa.

## Conclusion

Life expectancy at birth in Kenya, Uganda, and the United Republic of Tanzania progressively increased from 1960 to 2021. The trend dropped between 1970 and 2000 and between 2020, and 2021. High infant, maternal, and child mortality rates, and later, the high prevalence of HIV and AIDS, immunizable diseases, and the emergence of the COVID-19 pandemic, respectively are the likely reasons for the LE_0_ losses. Males and females in the three East African countries showed substantial life expectancy gaps in the last 61 years. Life expectancy losses in Uganda in the 70s were likely due to the political and economic turmoil during the brutal Amin regime and the other two countries in the 80s were likely due to HIV and AIDS and other immunizable diseases, and in 2020 and 2021 to COVID-19 pandemic. The COVID-19 pandemic contributed to life expectancy losses more in Kenya than United Republic of Tanzania and Uganda likely due to deaths related to the virus itself or the control measures. Even though many studies show life expectancy gains in Kenya, Uganda, and Tanzania over the 61 years, political stability, economic growth, health systems strengthening, control of infectious diseases, and epidemics were critical in the LE_0_ gains. However, a more comprehensive study is warranted to assess the actual impact of the public health interventions in LE_0_ gains in the three East African countries.

## Data Availability

All datasets supporting this article's conclusion are within this paper and are accessible by a reasonable request to the corresponding author.

https://www.google.com/search?q=united+nations+world+population+prospects+(wpps)&oq=United+Nations+World+Population+Prospects+(WPPs&gs_lcrp=EgZjaHJvbWUqBwgBECEYoAEyBggAEEUYOTIHCAEQIRigATIHCAIQIRigAdIBCjM4MzA1ajBqMTWoAgiwAgE&sourceid=chrome&ie=UTF-8

## Abbreviations

AIDS: Acquired Immune deficiency syndrome
ART: Antiretroviral therapy
CDC: United States Centres for Disease control and Prevention
COVID-19: Coronavirus disease-19
HIV: Human Immune Virus
LE_0_: Life expectancy at birth
MDGs: Millenium Development Goals
SARS-CoV-2: Severe acute respiratory coronavirus-2
SDGs: Sustainable Development Goals
WHO: World Health Organization
WPPs: World Population Prospects

## Declarations

### Ethics approval and consent to participate

This study on life expectancy on the population of Kenya, Uganda, and the United Republic of Tanzania was obtained from a public repository, and we conducted secondary data analysis. In addition, the study followed all relevant institutional guidelines and regulations on managing open data sources.

### Consent to publish

Not applicable.

### Authors’ contributions

DLK, GB, and JA designed this study. JA, GB, and DLK supervised data management. JA, GB, EO, and DLK analyzed and interpreted the data. JA, GB, EO, and DLK wrote and revised the manuscript. All Authors approved the manuscript.

### Authors’ Information

David Lagoro Kitara (DLK) is a Takemi fellow of Harvard University and a Professor at Gulu University, Faculty of Medicine, Department of Surgery, Gulu City, Uganda; Dr. Gaye Bamba (GB) is a founder member of the African Research Network (ARN), Dakar, Senegal; Joelle Abi Abboud (JA) is Research manager at the African Research Network (ARN), Paris, France; Dr. Emmanuel Olal is a physician and a public health specialist at Yotkom Medical Centre in Kitgum Municipality, Uganda.

## Acknowledgment

We thank the assistance from WPPs for the datasets obtained and Joelle Abi Abboud for a comprehensive data analysis for this study.

## References

1. Emily Oster, Ira Shoulson and E. Ray Dorsey. Limited Life Expectancy, Human Capital and Health Investments. The American Economic Review. 2013;5(103):1977–2002.

2. Gary S Becker, Tomas J Philipson, Rodrigo R. Soares. The Quantity and Quality of Life and the Evolution of World Inequality. Ameri can Economi c Revi ew. 2005; 95(1): 277–291.

3. Kevin M Murphy, Robert H Topher. The value of health and longevity. National Bureau of Economic Research. 2005. Working Paper 11405. doi 10.3386/w11405.

4. Laura Leker, Grégory Ponthière. Education, Life Expectancy and Family Bargaining: The Ben-Porath Effect Revisited. 2012. ffhalshs-00715104.

5. Michael D. Hurd, Kathleen Mcgarry. The Predictive Validity of Subjective Probabilities of Survival. The Economic Journal. 2002;112(482):966–985. DOI:10.1111/1468-0297.00065

6. Delavande A, Gine X, McKenzie D. Measuring subjective expectations in developing countries: A critical review and new evidence. Journal of development economics. 2011;94:151–163.

7. Delavande A, Rohwedder S. Differential survival in Europe and the United States: Estimates based on subjective probabilities of survival. Demography. 2011;48:1377–1400.

8. UNAIDS. HIV treatment in Africa: A looming crisis. Geneva: UNAIDS; 2015.

9. World Bank (WB). Life expectancy at birth, total years. 2010. https://data.worldbank.org/indicator/SP.DYN.LE00.IN

10. United Nations Development Population (UNDP). FACT SHEET: Africa Human Development Report 2016. Accelerating gender equality and women’s empowerment in Africa. 016. https://www.undp.org/sites/g/files/zskgke326/files/publications/AfHDR_FactSheetsD%20EN_web.pdf

11. UNAIDS and WHO. Ambitious treatment targets: Writing the final chapter of AIDS Epidemics. 2011. https://www.unaids.org/sites/default/files/media_asset/JC2670_UNAIDS_Treatment_Targ ets_en.pdf

12. Guisan Mc and Exposito P. Life Expectancy, Education And Development In African Countries 1980-2014: Improvements And International Comparisons. Applied Econometrics and International Development.2016:16–2.

13. Catherine Mcauliffe, Sweta C Saxena, and Masafumi Yabara. Sustaining Growth in the East African Community. International Monetary Fund.11-38 https://www.google.com/search?q=Economic+growth+in+the+East+African+region+over+the+last+60+years&oq=Economic+growth+in+the+East+African+region+over+the+last+60+years&gs_lcrp=EgZjaHJvbWUyBggAEEUYOdIBCjI1MjMzajBqMTWoAgCwAgA&sourceid=chrome&ie=UTF-8.

14. OCHA services. World Population Prospects 2022: Summary of Results. eliefweb. 2022. https://reliefweb.int/report/world/world-population-prospects-2022-summary-results?

15. World Population Prospects - Population Division - United Nations [Internet]. [cited 2023 Aug 1]. Available from: https://population.un.org/wpp/Download/Standard/Mortality/.

16. Softonic [Internet]. [cited 2023 Aug 7]. Download RStudio Desktop - free - latest version. Available from: https://rstudio-desktop.en.softonic.com.

17. Timothy C, Jacqueline N Milton. Basic Statistical Analysis Using the R Statistical Package. RStudio Team. 2021. https://sphweb.bumc.bu.edu/otlt/MPH-Modules/BS/R/R-Manual/R-Manual_print.html

18. World Health Organization (WHO). GHE: Life expectancy and healthy life expectancy. 2021. https://www.who.int/data/gho/data/themes/mortality-and-global-health-estimates/ghe-life-expectancy-and-healthy-life-expectancy

19. Muhammad Zakir Hossin. The male disadvantage in life expectancy: can we close the gender gap? International Health. 2021;13(5):482–484.

20. Zarulli V, Barthold Jones JA, Oksuzyan A, Lindahl-Jacobsen R, Christensen K, Vaupel JW. Women live longer than men even during severe famines and epidemics. Proc Natl Acad Sci U S A. 2018;115(4):E832-E840.doi: 10.1073/pnas.1701535115.

21. Lori Youmshajekian. Why the life expecytancy gap between men and women is growing. Scientific American. 2023. https://www.scientificamerican.com/article/why-the-life-expectancy-gap-between-men-and-women-is-growing/

22. Virginia Zarulli, Julia A Barthold Jonesa, Anna Oksuzyan, Rune Lindahl-Jacobsen, Kaare Christensena, and James W. Vaupela Women live longer than men even during severe famines and epidemics. PNAS. 2018;115(4):E832-E840. 10.1073/pnas.1701535115.

23. Worldometer. Life expectancy of the world population. 2020. Available from: https://www.worldometers.info/demographics/life-expectancy [accessed 25 July 2020].

24. Austad SN. Why women live longer than men: sex differences in longevity. Gend Med. 2006;3(4):79–92.

25. Regan JC, Partridge L. Gender and longevity: why do men die earlier than women? Comparative and experimental evidence. Best Pract Res Clin Endocrinol Metab. 2013;27(4):467–79

26. Austad SN, Fischer KE. Sex differences in lifespan. Cell Metab. 2016;23(6):1022–33.

27. Luy M, Minagawa Y. Gender gaps – life expectancy and proportion of life in poor health. Health Rep. 2014;25(12):12–19.

28. Eriksson JG, Kajantie E, Osmond C. Boys live dangerously in the womb. Am J Hum Biol. 2010;22(3):330–5.

29. Glover V. Maternal stress during pregnancy and infant and child outcomes. In:Wenzel A, editor. The Oxford handbook of perinatal psychology. Oxford: Oxford University Press; 2016:268–83.

30. Ingemarsson I. Gender aspects of preterm birth. BJOG. 2003;110(Suppl 20):34–8.

31. Marais GAB, Gaillard J, Vieira C. Sex gap in aging and longevity□: can sex chromosomes play a role? Biol Sex Differ. 2018;9(33):1–14.

32. Frank SA. Evolution: mitochondrial burden on male health. Curr Biol. 2012;22(18):R797–9.

33. African Development Bank Group (ADBG). Child mortality rates in Sub-Saharan Africa fall dramatically. African development fund. 014. https://www.afdb.org/fr/news-and-events/child-mortality-rates-in-sub-saharan-africa-fall-dramatically-13279#:~:text=Sub%2DSaharan%20Africa%20saw%20unprecedented,98%20per%201%2C000%20in%202012.

34. World Health Organization (WHO). What are the leading causes of mortality in the African Region. 2023. https://files.aho.afro.who.int/afahobckpcontainer/production/files/iAHO_Mortality_Regi onal-Factsheet.pdf

