## Supplementary material for "Comparative analysis of the evolution of Life Expectancy in the United Republic of Tanzania, Uganda, and Kenya in 61 years (1960-2021): A secondary data analysis of the World Population Prospects (WPPs) on the three East African countries": Table 1

```
Table 1: LE in Kenya from 1960 t0 2021 (61 years)
 Single term deletions
 Model: LE ~ Year + age + sexe
        Df Sum of Sq     RSS   AIC  Pr(>Chi)
 <none>               129497 29384
 Year   61     10753  140250 30261 <2.2e-16***
 age     1   4269151 4398648 73534 <2.2e-16***
 sexe    1     10870  140367 30392 <2.2e-16***
 Call: lm(formula = LE ~ Year + age + sexe, data = df)
 Residuals:
     Min      1Q     Median      3Q     Max
   -7.8444 -2.7652 -0.7726    2.3305   10.4877
 Coefficients:
               Estimate Std. Error  t value  Pr(>|t|)
 (Intercept) 56.9976620  0.2339253  243.658  <2e-16***
 Year1961     0.1814851  0.3207818    0.566 0.571568
 Year1962     0.3023267  0.3207818    0.942 0.345971
 Year1963     0.4063366  0.3207818    1.267 0.205284
 Year1964     0.5110891  0.3207818    1.593 0.111127
 Year1965     0.5744554  0.3207818    1.791 0.073350 .
 Year1966     0.6798020  0.3207818    2.119 0.034093 *
 Year1967     0.7621782  0.3207818    2.376 0.017516 *
 Year1968     0.8365347  0.3207818    2.608 0.009123 **
 Year1969     0.9397525  0.3207818    2.930 0.003400 **
 Year1970     1.0463366  0.3207818    3.262 0.001110 **
 Year1971     1.3341584  0.3207818    4.159 3.22e-05 ***
 Year1972     1.6394059  0.3207818    5.111 3.26e-07 ***
 Year1973     1.8566832  0.3207818    5.788 7.29e-09 ***
 Year1974     1.6470792  0.3207818    5.135 2.87e-07 ***
 Year1975     1.5241089  0.3207818    4.751 2.04e-06 ***
 Year1976     1.3725248  0.3207818    4.279 1.89e-05 ***
 Year1977     1.5393564  0.3207818    4.799 1.61e-06 ***
 Year1978     1.6054455  0.3207818    5.005 5.67e-07 ***
 Year1979     1.8739109  0.3207818    5.842 5.30e-09 ***
 Year1980     2.1529703  0.3207818    6.712 2.01e-11 ***
 Year1981     3.6876238  0.3207818   11.496  < 2e-16 ***
 Year1982     3.3621287  0.3207818   10.481  < 2e-16 ***
 Year1983     3.2323762  0.3207818   10.077  < 2e-16 ***
 Year1984     2.9826238  0.3207818    9.298  < 2e-16 ***
 Year1985     2.8269307  0.3207818    8.813  < 2e-16 ***
 Year1986     2.6147525  0.3207818    8.151 3.95e-16 ***
 Year1987     2.5348020  0.3207818    7.902 2.98e-15 ***
 Year1988     2.3183168  0.3207818    7.227 5.22e-13 ***
 Year1989     2.1445050  0.3207818    6.685 2.40e-11 ***
 Year1990     2.0347030  0.3207818    6.343 2.33e-10 ***
 Year1991     1.8455941  0.3207818    5.753 8.95e-09 ***
 Year1992     1.5829703  0.3207818    4.935 8.13e-07 ***
 Year1993     1.3409406  0.3207818    4.180 2.93e-05 ***
 Year1994     1.2180198  0.3207818    3.797 0.000147 ***
 Year1995     0.8829208  0.3207818    2.752 0.005925 **
 Year1996     0.6202475  0.3207818    1.934 0.053191 .
 Year1997     0.3854455  0.3207818    1.202 0.229548
 Year1998     0.1556931  0.3207818    0.485 0.627433
 Year1999     0.0122277  0.3207818    0.038 0.969594
 Year2000    -0.1610396  0.3207818   -0.502 0.615661
 Year2001    -0.2449505  0.3207818   -0.764 0.445117
 Year2002    -0.1044554  0.3207818   -0.326 0.744712
 Year2003     0.0600990  0.3207818    0.187 0.851388
 Year2004     0.2977228  0.3207818    0.928 0.353365
 Year2005     0.6496040  0.3207818    2.025 0.042882 *
 Year2006     0.9680693  0.3207818    3.018 0.002551 **
 Year2007     1.1701485  0.3207818    3.648 0.000266 ***
 Year2008     1.3562871  0.3207818    4.228 2.37e-05 ***
 Year2009     1.5622772  0.3207818    4.870 1.13e-06 ***
 Year2010     1.6330198  0.3207818    5.091 3.62e-07 ***
 Year2011     1.7584653  0.3207818    5.482 4.29e-08 ***
 Year2012     1.7350990  0.3207818    5.409 6.46e-08 ***
 Year2013     1.8065347  0.3207818    5.632 1.82e-08 ***
 Year2014     1.9940594  0.3207818    6.216 5.25e-10 ***
 Year2015     1.9949505  0.3207818    6.219 5.16e-10 ***
 Year2016     2.0813861  0.3207818    6.488 9.00e-11 ***
 Year2017     2.2208416  0.3207818    6.923 4.63e-12 ***
 Year2018     2.2973762  0.3207818    7.162 8.41e-13 ***
 Year2019     2.4152475  0.3207818    7.529 5.45e-14 ***
 Year2020     2.2437129  0.3207818    6.995 2.80e-12 ***
 Year2021     1.1619307  0.3207818    3.622 0.000293 ***
 age         -0.6332713  0.0009881 -640.915  < 2e-16 ***
 sexeMale    -1.8632482  0.0576141  -32.340  < 2e-16 ***
 Signif. codes: 0'***' 0.001'**' 0.01'*' 0.05 '.' 0.1 '1'
 Residual standard error: 3.224 on 12460 degrees of freedom
 Multiple R-squared:  0.9707, Adjusted R-squared:  0.9706
 F-statistic:  6553 on 63 and 12460 DF,  p-value: <2.2e-16
```
