## Supplementary material for "Comparative analysis of the evolution of Life Expectancy in the United Republic of Tanzania, Uganda, and Kenya in 61 years (1960-2021): A secondary data analysis of the World Population Prospects (WPPs) on the three East African countries": Table 2

**Table 2: Analysis of Variance for LE_0_ in Kenya from 1960 to 2021.**
 **Response: LE**
 Df Sum Sq Mean Sq	F value	Pr(>F)
 Year	61	10753	176	16.961	<2.2e-16***
 age	1 4269151 4269151 410771.897	<2.2e-16***
 sexe	1	10870	10870	1045.884	<2.2e-16***
 **Residuals:** 12460 129497	10
 **Call:** lm(formula = res1 ~ Year + age + sexe, data = df)
 **Residuals:**
 Min	1Q	Median	3Q	Max
 -7.8444 -2.7652 -0.7726	2.3305	10.4877
 **Coefficients:**
 Estimate Std. Error	t value Pr(>|t|)
 (Intercept) 3.010e-15 2.339e-01	0	1
 Year1961	1.557e-14 3.208e-01	0	1
 Year1962	-1.020e-14 3.208e-01	0	1
 Year1963	3.698e-14 3.208e-01	0	1
 Year1964	4.490e-14 3.208e-01	0	1
 Year1965	-5.268e-14 3.208e-01	0	1
 Year1966	5.491e-14 3.208e-01	0	1
 Year1967	-8.671e-14 3.208e-01	0	1
 Year1968	-3.015e-14 3.208e-01	0	1
 Year1969	7.618e-14 3.208e-01	0	1
 Year1970	-1.760e-14 3.208e-01	0	1
 Year1971	-3.742e-14 3.208e-01	0	1
 Year1972	-6.666e-14 3.208e-01	0	1
 Year1973	-2.238e-14 3.208e-01	0	1
 Year1974	1.358e-14 3.208e-01	0	1
 Year1975	2.626e-14 3.208e-01	0	1
 Year1976	-4.129e-15 3.208e-01	0	1
 Year1977	-2.842e-14 3.208e-01	0	1
 Year1978	-2.197e-14 3.208e-01	0	1
 Year1979	6.309e-15 3.208e-01	0	1
 Year1980	-2.478e-15 3.208e-01	0	1
 Year1981	-1.724e-17 3.208e-01	0	1
 Year1982	-5.762e-16 3.208e-01	0	1
 Year1983	2.307e-15 3.208e-01	0	1
 Year1984	2.508e-15 3.208e-01	0	1
 Year1985	2.398e-15 3.208e-01	0	1
 Year1986	4.456e-15 3.208e-01	0	1
 Year1987	-9.844e-15 3.208e-01	0	1
 Year1988	4.519e-15 3.208e-01	0	1
 Year1989	-5.632e-15 3.208e-01	0	1
 Year1990	-1.078e-14 3.208e-01	0	1
 Year1991	8.552e-16 3.208e-01	0	1
 Year1992	7.701e-15 3.208e-01	0	1
 Year1993	-1.589e-14 3.208e-01	0	1
 Year1994	1.363e-14 3.208e-01	0	1
 Year1995	1.575e-14 3.208e-01	0	1
 Year1996	-4.321e-15 3.208e-01	0	1
 Year1997	-1.040e-14 3.208e-01	0	1
 Year1998	-1.751e-14 3.208e-01	0	1
 Year1999	7.151e-16 3.208e-01	0	1
 Year2000	-6.912e-15 3.208e-01	0	1
 Year2001	1.481e-15 3.208e-01	0	1
 Year2002	-7.926e-15 3.208e-01	0	1
 Year2003	1.591e-14 3.208e-01	0	1
 Year2004	-1.804e-14 3.208e-01	0	1
 Year2005	1.231e-14 3.208e-01	0	1
 Year2006	-7.937e-15 3.208e-01	0	1
 Year2007	-5.725e-15 3.208e-01	0	1
 Year2008	-9.183e-15 3.208e-01	0	1
 Year2009	4.007e-15 3.208e-01	0	1
 Year2010	8.629e-15 3.208e-01	0	1
 Year2011	9.692e-15 3.208e-01	0	1
 Year2012	-3.618e-15 3.208e-01	0	1
 Year2013	8.610e-15 3.208e-01	0	1
 Year2014	5.311e-15 3.208e-01	0	1
 Year2015	1.465e-15 3.208e-01	0	1
 Year2016	-2.821e-15 3.208e-01	0	1
 Year2017	-5.469e-15 3.208e-01	0	1
 Year2018	1.942e-15 3.208e-01	0	1
 Year2019	-2.226e-16 3.208e-01	0	1
 Year2020	-1.016e-15 3.208e-01	0	1
 Year2021	-2.033e-15 3.208e-01	0	1
 age	-2.858e-17 9.881e-04	0	1
 sexeMale	1.349e-16 5.761e-02	0	1
 **Signif. codes:** 0'***' 0.001'**' 0.01'*' 0.05'.' 0.1 '1'

```
 Residual standard error:3.224 on 12460 degrees of freedom
 Multiple R-squared:5.551e-29,  Adjusted R-squared:-0.005056
 F-statistic: 1.098e-26 on 63 and 12460 DF,  p-value: 1
```
