## Supplementary material for "Comparative analysis of the evolution of Life Expectancy in the United Republic of Tanzania, Uganda, and Kenya in 61 years (1960-2021): A secondary data analysis of the World Population Prospects (WPPs) on the three East African countries": Table 3

**Table 3: LE_0_ analysis of Uganda from 1960 to 2021 (Using 1960 as reference year).**

```
Single term deletions
Model: LE ~ Year + age + sexe
        Df Sum of Sq     RSS   AIC     Pr(>Chi)
 <none>               139830 30346
 Year   61     17143  156973 31672   <2.2e-16 ***
 age     1   3676585 3816415 71756   <2.2e-16 ***
 sexe    1     18865  158695 31929   <2.2e-16 ***
 Call:lm(formula = LE ~ Year + age + sexe, data = df)
 Residuals:
      Min       1Q   Median       3Q        Max
  -11.8694   -2.5411  -0.9172    2.1930    10.6407
 Coefficients:
              Estimate Std. Error  t value Pr(>|t|)
 (Intercept) 54.353975   0.243079  223.606  < 2e-16 ***
 Year1961     0.304901   0.333335    0.915 0.360368
 Year1962     0.588020   0.333335    1.764 0.077748 .
 Year1963     0.709356   0.333335    2.128 0.033352 *
 Year1964     0.836040   0.333335    2.508 0.012151 *
 Year1965     0.906535   0.333335    2.720 0.006545 **
 Year1966     0.852426   0.333335    2.557 0.010562 *
 Year1967     1.031485   0.333335    3.094 0.001976 **
 Year1968     1.051782   0.333335    3.155 0.001607 **
 Year1969     1.117574   0.333335    3.353 0.000803 ***
 Year1970     1.112178   0.333335    3.337 0.000851 ***
 Year1971     0.017624   0.333335    0.053 0.957836
 Year1972     0.049901   0.333335    0.150 0.881002
 Year1973     0.133564   0.333335    0.401 0.688654
 Year1974     0.005941   0.333335    0.018 0.985781
 Year1975     0.027129   0.333335    0.081 0.935136
 Year1976    -0.064356   0.333335   -0.193 0.846909
 Year1977    -0.122921   0.333335   -0.369 0.712312
 Year1978    -0.249059   0.333335   -0.747 0.454972
 Year1979    -0.311535   0.333335   -0.935 0.350013
 Year1980    -0.959950   0.333335   -2.880 0.003986 **
 Year1981    -0.992079   0.333335   -2.976 0.002924 **
 Year1982    -1.328861   0.333335   -3.987 6.74e-05 ***
 Year1983    -1.396386   0.333335   -4.189 2.82e-05 ***
 Year1984    -1.529455   0.333335   -4.588 4.51e-06 ***
 Year1985    -1.059109   0.333335   -3.177 0.001490 **
 Year1986    -1.242624   0.333335   -3.728 0.000194 ***
 Year1987    -0.823564   0.333335   -2.471 0.013499 *
 Year1988    -0.891980   0.333335   -2.676 0.007462 **
 Year1989    -1.142574   0.333335   -3.428 0.000611 ***
 Year1990    -1.316139   0.333335   -3.948 7.91e-05 ***
 Year1991    -1.571040   0.333335   -4.713 2.47e-06 ***
 Year1992    -1.720990   0.333335   -5.163 2.47e-07 ***
 Year1993    -1.956535   0.333335   -5.870 4.48e-09 ***
 Year1994    -2.001980   0.333335   -6.006 1.96e-09 ***
 Year1995    -1.963911   0.333335   -5.892 3.92e-09 ***
 Year1996    -1.972822   0.333335   -5.918 3.34e-09 ***
 Year1997    -1.908713   0.333335   -5.726 1.05e-08 ***
 Year1998    -1.915099   0.333335   -5.745 9.39e-09 ***
 Year1999    -1.784208   0.333335   -5.353 8.82e-08 ***
 Year2000    -1.703861   0.333335   -5.112 3.24e-07 ***
 Year2001    -1.621188   0.333335   -4.864 1.17e-06 ***
 Year2002    -1.532277   0.333335   -4.597 4.33e-06 ***
 Year2003    -1.440000   0.333335   -4.320 1.57e-05 ***
 Year2004    -1.143861   0.333335   -3.432 0.000602 ***
 Year2005    -0.695396   0.333335   -2.086 0.036983 *
 Year2006    -0.259059   0.333335   -0.777 0.437071
 Year2007    -0.069208   0.333335   -0.208 0.835527
 Year2008     0.017426   0.333335    0.052 0.958309
 Year2009     0.152030   0.333335    0.456 0.648335
 Year2010     0.247129   0.333335    0.741 0.458475
 Year2011     0.514752   0.333335    1.544 0.122553
 Year2012     0.762673   0.333335    2.288 0.022154 *
 Year2013     1.040248   0.333335    3.121 0.001808 **
 Year2014     1.191535   0.333335    3.575 0.000352 ***
 Year2015     1.375495   0.333335    4.126 3.71e-05 ***
 Year2016     1.521881   0.333335    4.566 5.03e-06 ***
 Year2017     1.647723   0.333335    4.943 7.79e-07 ***
 Year2018     1.846535   0.333335    5.540 3.09e-08 ***
 Year2019     1.902228   0.333335    5.707 1.18e-08 ***
 Year2020     1.634901   0.333335    4.905 9.48e-07 ***
 Year2021     1.344802   0.333335    4.034 5.51e-05 ***
 age         -0.587681   0.001027 -572.375  < 2e-16 ***
 sexeMale    -2.454650   0.059869  -41.001  < 2e-16 ***
 Signif. codes:0'***' 0.001'**' 0.01'*' 0.05'.' 0.1'1'
 Residual standard error: 3.35 on 12460 degrees of freedom
 Multiple R-squared:0.9637, Adjusted R-squared:0.9635
 F-statistic:5251 on 63 and 12460 DF,  p-value: <2.2e-16
```
