## Supplementary material for "Comparative analysis of the evolution of Life Expectancy in the United Republic of Tanzania, Uganda, and Kenya in 61 years (1960-2021): A secondary data analysis of the World Population Prospects (WPPs) on the three East African countries": Table 4

**Table 2:Analysis of Variance LE_0_ in Uganda from 1960-2021.**
 **Response: LE**
 Df Sum Sq Mean Sq	F value	Pr(>F)
 Year	61	17143	281	25.043	<2.2e-16***
 age	1 3676585 3676585 327613.374	<2.2e-16***
 sexe	1	18865	18865	1681.045	<2.2e-16***
 Residuals 12460 139830	11
 **Call:** lm(formula = res1 ~ Year + age + sexe, data = df)
 **Residuals:**
 Min	1Q	Median	3Q	Max
 -11.8694 -2.5411 -0.9172	2.1930	10.6407
 **Coefficients:**
 Estimate	Std. Error t value Pr(>|t|)
 (Intercept) 6.850e-15 2.431e-01	0	1
 Year1961	1.436e-14 3.333e-01	0	1
 Year1962	-1.224e-14 3.333e-01	0	1
 Year1963	3.598e-14 3.333e-01	0	1
 Year1964	4.323e-14 3.333e-01	0	1
 Year1965	-6.139e-14 3.333e-01	0	1
 Year1966	5.806e-14 3.333e-01	0	1
 Year1967	-9.997e-14 3.333e-01	0	1
 Year1968	-3.582e-14 3.333e-01	0	1
 Year1969	8.491e-14 3.333e-01	0	1
 Year1970	-2.262e-14 3.333e-01	0	1
 Year1971	-4.503e-14 3.333e-01	0	1
 Year1972	-8.045e-14 3.333e-01	0	1
 Year1973	-2.915e-14 3.333e-01	0	1
 Year1974	1.450e-14 3.333e-01	0	1
 Year1975	2.527e-14 3.333e-01	0	1
 Year1976	-8.239e-15 3.333e-01	0	1
 Year1977	-3.624e-14 3.333e-01	0	1
 Year1978	-2.800e-14 3.333e-01	0	1
 Year1979	6.550e-15 3.333e-01	0	1
 Year1980	-7.156e-15 3.333e-01	0	1
 Year1981	-3.262e-15 3.333e-01	0	1
 Year1982	-4.514e-15 3.333e-01	0	1
 Year1983	-2.413e-15 3.333e-01	0	1
 Year1984	-8.959e-16 3.333e-01	0	1
 Year1985	-5.599e-16 3.333e-01	0	1
 Year1986	-1.056e-15 3.333e-01	0	1
 Year1987	-1.151e-14 3.333e-01	0	1
 Year1988	1.653e-15 3.333e-01	0	1
 Year1989	-7.691e-15 3.333e-01	0	1
 Year1990	-1.358e-14 3.333e-01	0	1
 Year1991	-2.802e-15 3.333e-01	0	1
 Year1992	5.282e-15 3.333e-01	0	1
 Year1993	-1.646e-14 3.333e-01	0	1
 Year1994	8.709e-15 3.333e-01	0	1
 Year1995	1.033e-14 3.333e-01	0	1
 Year1996	-7.621e-15 3.333e-01	0	1
 Year1997	-1.309e-14 3.333e-01	0	1
 Year1998	-1.885e-14 3.333e-01	0	1
 Year1999	-3.245e-15 3.333e-01	0	1
 Year2000	-9.174e-15 3.333e-01	0	1
 Year2001	-1.633e-15 3.333e-01	0	1
 Year2002	-9.686e-15 3.333e-01	0	1
 Year2003	9.601e-15 3.333e-01	0	1
 Year2004	-1.978e-14 3.333e-01	0	1
 Year2005	7.369e-15 3.333e-01	0	1
 Year2006	-9.887e-15 3.333e-01	0	1
 Year2007	-7.826e-15 3.333e-01	0	1
 Year2008	-1.326e-14 3.333e-01	0	1
 Year2009	-1.508e-16 3.333e-01	0	1
 Year2010	2.701e-15 3.333e-01	0	1
 Year2011	5.524e-15 3.333e-01	0	1
 Year2012	-7.247e-15 3.333e-01	0	1
 Year2013	5.855e-15 3.333e-01	0	1
 Year2014	2.477e-15 3.333e-01	0	1
 Year2015	-2.522e-15 3.333e-01	0	1
 Year2016	-4.028e-15 3.333e-01	0	1
 Year2017	-8.505e-15 3.333e-01	0	1
 Year2018	-1.211e-15 3.333e-01	0	1
 Year2019	-9.259e-16 3.333e-01	0	1
 Year2020	-1.106e-16 3.333e-01	0	1
 Year2021	-4.507e-15 3.333e-01	0	1
 age	-2.722e-17 1.027e-03	0	1
 sexeMale	5.238e-16 5.987e-02	0	1
 **Signif. codes:**0'***' 0.001'**' 0.01'*' 0.05'.' 0.1 '1'

```
 Residual standard error:3.35 on 12460 degrees of freedom
 Multiple R-squared: 6.26e-29,   Adjusted R-squared:-0.005056
 F-statistic: 1.238e-26 on 63 and 12460 DF,  p-value: 1
```
