## Supplementary material for "Comparative analysis of the evolution of Life Expectancy in the United Republic of Tanzania, Uganda, and Kenya in 61 years (1960-2021): A secondary data analysis of the World Population Prospects (WPPs) on the three East African countries": Table 5

**Table 5: LE_0_ in the United Republic of Tanzania from 1960 to 2021.**

```
 Single term deletions
 Model:LE ~ Year + age + sexe
        Df Sum of Sq     RSS   AIC  Pr(>Chi)
 <none>               135478 29950
 Year   61     32147  167624 32494  <2.2e-16***
 age     1   4043394 4178871 72892  <2.2e-16***
 sexe    1     14021  149499 31181  <2.2e-16***
 Call:lm(formula = LE ~ Year + age + sexe, data = df)
 Residuals:
      Min       1Q     Median       3Q        Max
   -10.5477  -2.5771  -0.8319     2.0682    11.1222
 Coefficients:
             Estimate  Std. Error  t value   Pr(>|t|)
 (Intercept) 54.353916   0.239266  227.169  <2e-16***
 Year1961     0.047178   0.328106    0.144 0.885669
 Year1962     0.104208   0.328106    0.318 0.750790
 Year1963     0.159406   0.328106    0.486 0.627091
 Year1964     0.232277   0.328106    0.708 0.479000
 Year1965     0.309010   0.328106    0.942 0.346314
 Year1966     0.426733   0.328106    1.301 0.193421
 Year1967     0.553416   0.328106    1.687 0.091686 .
 Year1968     0.820198   0.328106    2.500 0.012439 *
 Year1969     1.076287   0.328106    3.280 0.001040 **
 Year1970     1.242426   0.328106    3.787 0.000153 ***
 Year1971     1.402475   0.328106    4.274 1.93e-05 ***
 Year1972     1.496881   0.328106    4.562 5.11e-06 ***
 Year1973     1.774356   0.328106    5.408 6.50e-08 ***
 Year1974     1.807129   0.328106    5.508 3.71e-08 ***
 Year1975     1.882178   0.328106    5.736 9.89e-09 ***
 Year1976     2.026832   0.328106    6.177 6.72e-10 ***
 Year1977     2.236733   0.328106    6.817 9.72e-12 ***
 Year1978     2.390297   0.328106    7.285 3.41e-13 ***
 Year1979     2.609010   0.328106    7.952 2.00e-15 ***
 Year1980     2.722426   0.328106    8.297  < 2e-16 ***
 Year1981     2.879703   0.328106    8.777  < 2e-16 ***
 Year1982     2.917673   0.328106    8.892  < 2e-16 ***
 Year1983     2.947277   0.328106    8.983  < 2e-16 ***
 Year1984     2.974653   0.328106    9.066  < 2e-16 ***
 Year1985     2.954356   0.328106    9.004  < 2e-16 ***
 Year1986     2.958267   0.328106    9.016  < 2e-16 ***
 Year1987     2.901733   0.328106    8.844  < 2e-16 ***
 Year1988     2.870990   0.328106    8.750  < 2e-16 ***
 Year1989     2.704208   0.328106    8.242  < 2e-16 ***
 Year1990     2.461931   0.328106    7.503 6.64e-14 ***
 Year1991     2.298267   0.328106    7.005 2.60e-12 ***
 Year1992     2.084653   0.328106    6.354 2.18e-10 ***
 Year1993     1.921287   0.328106    5.856 4.87e-09 ***
 Year1994     1.820297   0.328106    5.548 2.95e-08 ***
 Year1995     1.719208   0.328106    5.240 1.63e-07 ***
 Year1996     1.578267   0.328106    4.810 1.53e-06 ***
 Year1997     1.385495   0.328106    4.223 2.43e-05 ***
 Year1998     1.328762   0.328106    4.050 5.16e-05 ***
 Year1999     1.571386   0.328106    4.789 1.69e-06 ***
 Year2000     1.631931   0.328106    4.974 6.65e-07 ***
 Year2001     1.773713   0.328106    5.406 6.57e-08 ***
 Year2002     1.860149   0.328106    5.669 1.47e-08 ***
 Year2003     2.130297   0.328106    6.493 8.75e-11 ***
 Year2004     2.265594   0.328106    6.905 5.26e-12 ***
 Year2005     2.415941   0.328106    7.363 1.91e-13 ***
 Year2006     2.558762   0.328106    7.799 6.76e-15 ***
 Year2007     2.703614   0.328106    8.240  < 2e-16 ***
 Year2008     2.858960   0.328106    8.714  < 2e-16 ***
 Year2009     3.122129   0.328106    9.516  < 2e-16 ***
 Year2010     3.630297   0.328106   11.064  < 2e-16 ***
 Year2011     3.998317   0.328106   12.186  < 2e-16 ***
 Year2012     4.408317   0.328106   13.436  < 2e-16 ***
 Year2013     4.769901   0.328106   14.538  < 2e-16 ***
 Year2014     5.129356   0.328106   15.633  < 2e-16 ***
 Year2015     5.462030   0.328106   16.647  < 2e-16 ***
 Year2016     5.751287   0.328106   17.529  < 2e-16 ***
 Year2017     5.986386   0.328106   18.245  < 2e-16 ***
 Year2018     6.170297   0.328106   18.806  < 2e-16 ***
 Year2019     6.369851   0.328106   19.414  < 2e-16 ***
 Year2020     5.647970   0.328106   17.214  < 2e-16 ***
 Year2021     5.372871   0.328106   16.375  < 2e-16 ***
 age         -0.616300   0.001011 -609.815  < 2e-16 ***
 sexeMale    -2.116171   0.058930  -35.910  < 2e-16 ***
 Signif. codes:0'***' 0.001'**' 0.01'*' 0.05'.' 0.1'1'
 Residual standard error: 3.297 on 12460 degrees of freedom
 Multiple R-squared:0.9679, Adjusted R-squared:0.9678
 F-statistic: 5970 on 63 and 12460 DF, p-value:<2.2e-16
```
