## Supplementary material for "Comparative analysis of the evolution of Life Expectancy in the United Republic of Tanzania, Uganda, and Kenya in 61 years (1960-2021): A secondary data analysis of the World Population Prospects (WPPs) on the three East African countries": Table 6

```
Table 6:Analysis of Variance for United Republic of Tanzania (1960-2021).
 Response:LE
              Df  Sum Sq  Mean Sq    F value    Pr(>F)
 Year         61   32147     527     48.468  <2.2e-16***
 age           1 4043394 4043394 371874.598  <2.2e-16***
 sexe          1   14021   14021   1289.540  <2.2e-16***
 Residuals: 12460  135478      11
 Call: lm(formula = res1 ~ Year + age + sexe, data = df)
 Residuals:
      Min       1Q    Median       3Q        Max
  -10.5477  -2.5771  -0.8319     2.0682    11.1222
 Coefficients:
               Estimate Std. Error   t value Pr(>|t|)
 (Intercept) -1.124e-14  2.393e-01       0        1
 Year1961     4.098e-14  3.281e-01       0        1
 Year1962     6.252e-14  3.281e-01       0        1
 Year1963     3.677e-15  3.281e-01       0        1
 Year1964     2.346e-14  3.281e-01       0        1
 Year1965    -1.668e-14  3.281e-01       0        1
 Year1966     3.890e-14  3.281e-01       0        1
 Year1967    -4.096e-14  3.281e-01       0        1
 Year1968    -8.090e-15  3.281e-01       0        1
 Year1969     5.315e-14  3.281e-01       0        1
 Year1970    -1.052e-15  3.281e-01       0        1
 Year1971    -1.100e-14  3.281e-01       0        1
 Year1972    -2.404e-14  3.281e-01       0        1
 Year1973    -2.311e-15  3.281e-01       0        1
 Year1974     1.428e-14  3.281e-01       0        1
 Year1975     1.517e-14  3.281e-01       0        1
 Year1976     7.503e-15  3.281e-01       0        1
 Year1977     8.583e-15  3.281e-01       0        1
 Year1978     1.506e-14  3.281e-01       0        1
 Year1979     1.969e-15  3.281e-01       0        1
 Year1980     1.187e-14  3.281e-01       0        1
 Year1981     6.137e-15  3.281e-01       0        1
 Year1982     2.218e-14  3.281e-01       0        1
 Year1983     2.224e-14  3.281e-01       0        1
 Year1984     1.913e-14  3.281e-01       0        1
 Year1985     1.592e-14  3.281e-01       0        1
 Year1986     1.643e-14  3.281e-01       0        1
 Year1987    -7.608e-15  3.281e-01       0        1
 Year1988     1.653e-14  3.281e-01       0        1
 Year1989    -1.996e-16  3.281e-01       0        1
 Year1990    -5.392e-15  3.281e-01       0        1
 Year1991     8.618e-15  3.281e-01       0        1
 Year1992     1.927e-14  3.281e-01       0        1
 Year1993    -8.119e-15  3.281e-01       0        1
 Year1994     2.082e-14  3.281e-01       0        1
 Year1995     2.448e-14  3.281e-01       0        1
 Year1996     3.433e-15  3.281e-01       0        1
 Year1997    -1.765e-15  3.281e-01       0        1
 Year1998    -7.932e-15  3.281e-01       0        1
 Year1999     8.311e-15  3.281e-01       0        1
 Year2000     2.817e-15  3.281e-01       0        1
 Year2001     7.179e-15  3.281e-01       0        1
 Year2002     1.905e-15  3.281e-01       0        1
 Year2003     1.998e-14  3.281e-01       0        1
 Year2004    -5.864e-15  3.281e-01       0        1
 Year2005     1.461e-14  3.281e-01       0        1
 Year2006     4.044e-15  3.281e-01       0        1
 Year2007     3.712e-15  3.281e-01       0        1
 Year2008    -1.414e-15  3.281e-01       0        1
 Year2009     9.696e-15  3.281e-01       0        1
 Year2010     1.177e-14  3.281e-01       0        1
 Year2011     1.506e-14  3.281e-01       0        1
 Year2012     4.016e-15  3.281e-01       0        1
 Year2013     1.250e-14  3.281e-01       0        1
 Year2014     1.148e-14  3.281e-01       0        1
 Year2015     7.367e-15  3.281e-01       0        1
 Year2016     5.531e-15  3.281e-01       0        1
 Year2017     5.802e-15  3.281e-01       0        1
 Year2018     8.259e-15  3.281e-01       0        1
 Year2019     8.017e-15  3.281e-01       0        1
 Year2020     5.700e-15  3.281e-01       0        1
 Year2021     9.258e-15  3.281e-01       0        1
 age          5.070e-17  1.011e-03       0        1
 sexeMale    -5.119e-16  5.893e-02       0        1
 Residual standard error:3.297 on 12460 degrees of freedom
 Multiple R-squared:2.324e-29, Adjusted R-squared:-0.005056
 F-statistic: 4.596e-27 on 63 and 12460 DF, p-value: 1
```
